# Quantifying Inequities in COVID-19 Vaccine Distribution Over Time by social vulnerability, race and ethnicity, and location: A Population-Level Analysis in St. Louis and Kansas City, Missouri

**DOI:** 10.1101/2022.06.13.22276312

**Authors:** Aaloke Mody, Cory Bradley, Salil Redkar, Branson Fox, Ingrid Eshun-Wilson, Matifadza G. Hlatshwayo, Anne Trolard, Khai Hoan Tram, Lindsey M. Filiatreau, Franda Thomas, Matt Haslam, George Turabelidze, Vetta Sanders-Thompson, William G. Powderly, Elvin H. Geng

**Affiliations:** Washington University School of Medicine, St. Louis, Missouri, USA; St. Louis City Department of Health, St. Louis, Missouri, USA; Institute for Public Health, Washington University in St. Louis, St. Louis, Missouri, USA; University of Washington School of Medicine, Seattle, WA, USA; Missouri Department of Health and Senior Services, Jefferson City and St Louis, Missouri, USA; Brown School of Social Work, Washington University in St. Louis, St. Louis, Missouri, USA

**Keywords:** COVID-19 vaccinations, Racial Disparities, Lorenz Curve, Inequity, structural racism, vaccine locations, social vulnerability index

## Abstract

**BACKGROUND:** Equity in vaccination coverage is a cornerstone to a successful public health response to COVID-19. To deepen understand of the extent to which vaccination coverage compared to initial strategies for equitable vaccination, we explore primary vaccine series and booster rollout over time and by race/ethnicity, social vulnerability, and geography.

**METHODS AND FINDINGS:** We analyzed data from the Missouri State Department of Health and Senior Services on all COVID-19 vaccinations administered across 7 counties in the St. Louis region and 4 counties in the Kansas City Region. We compared rates of receiving the primary COVID-19 vaccine series and boosters relative to time, race/ethnicity, zip code-level social vulnerability index (SVI), vaccine location type, and COVID-19 disease burden. We adapted a well-established tool for measuring inequity—the Lorenz curve—to quantify inequities in COVID-19 vaccination relative to these key metrics. Between 12/15/2020 and 2/15/2022, 1,762,508 individuals completed the primary series and 871,896 had received a booster. During early phases of the primary series rollout, Black and Hispanic individuals from high SVI zip codes were vaccinated at less than half the rate of White individuals, but rates increased over time until they were higher than rates in White individuals after June 2021; Asian individuals maintained high levels of vaccination throughout. Increasing vaccination rates in Black and Hispanic communities corresponded with periods when more vaccinations were offered at small community-based sites such as pharmacies rather than larger health systems and mass vaccination sites. Using Lorenz curves, zip codes in the quartile with the lowest rates of primary series completion accounted for 19.3%, 18.1%, 10.8%, and 8.8% of vaccinations but represented 25% of either the total population, cases, deaths, or population-level SVI, respectively. When tracking Gini coefficients, these disparities were greatest earlier during rollout, but improvements were slow and modest and vaccine disparities remained across all metrics even after one year. Patterns of disparities for boosters were similar but often of much greater magnitude during rollout in Fall 2021. Study limitations include inherent limitations in vaccine registry dataset such as missing and misclassified race/ethnicity and zip code variables and potential changes in zip code population sizes since census enumeration.

**CONCLUSIONS:** Racial inequity in the initial COVID-19 vaccination and booster rollout in two large U.S. metropolitan areas were apparent across racial/ethnic communities, across levels of social vulnerability, over time, and across types of vaccination administration sites. Disparities in receipt of the primary vaccine series attenuated over time during a period in which sites of vaccination administration diversified, but were recapitulated during booster rollout. These findings highlight how public health strategies from the outset must directly target these deeply embedded structural and systemic determinants of disparities and track equity metrics over time to avoid perpetuating inequities in health care access.

**AUTHOR SUMMARY:** *Why Was This Study Done?:*

- Equitable vaccine strategies are critical for the public health response to COVID-19, but there is limited understanding of how vaccination campaigns compared to different metrics for equity.
- Many initial approaches to vaccine allocation sought to acknowledge the known disparities in exposure risk, disease burden, needs, and access by formally considering social vulnerability or race/ethnicity in plans to prioritize vaccinations, but there is limited empirical evaluation of how actual primary vaccine series and subsequent booster efforts aligned with the initial goals set out for equity.
- We quantify COVID-19 vaccine-related inequities in receipt of the primary vaccine series and booster across key equity metrics including race/ethnicity, social vulnerability, location, and time using a novel application of Lorenz curves and Gini coefficients—tools from economics to measure inequalities—in the St. Louis and Kansas City regions of Missouri.

*What Did the Researchers Do and Find?:*

- We analyzed data from the Missouri State Department of Health and Senior Services on all COVID-19 vaccinations administered in the St. Louis region and Kansas City Regions. We compared rates of receiving the primary COVID-19 vaccine series and boosters relative to time, race/ethnicity, zip code-level social vulnerability index (SVI), vaccine location type, and COVID-19 disease burden. We adapted Lorenz curves and Gini coefficients to quantify the inequities in COVID-19 vaccination relative to these key metrics and examined how they changed over time.
- Black and Hispanic individuals from high SVI zip codes completed the primary series at less than half the rate of White individuals during early phases of the primary series rollout, but surpassed rates in White individuals after June 2021. These relative increases in primary series completion rates in Black and Hispanic communities corresponded to periods when vaccinations became more available at small community-based sites.
- Lorenz curves demonstrated that zip codes in the quartile with the lowest rates of primary series completion accounted for 19.3%, 18.1%, 10.8%, and 8.8% of vaccinations but represented 25% of either the total population, cases, deaths, or population-level SVI, respectively. Tracking Gini coefficients over time demonstrated that these disparities were greatest earlier during rollout, but only improved slowly and modestly over time.
- Patterns of disparities for boosters were similar but often of much greater magnitude that those seen with completion of the primary vaccine series. patterns of disparities were similar but often of greater magnitude during booster rollout in Fall 2021.

*What Do These Findings Mean?:*

- Vaccination coverage for both the primary series and boosters demonstrated substantial disparities across race/ethnicity, levels of social vulnerability, types of vaccine administration sites, and over time.
- Despite well-documented inequities for COVID-19 and need for equitable vaccine approaches, the strategies employed did not overcome deeply entrenched systemic inequities in health care and society.
- Public health strategies must proactively target these deeply embedded structural determinants of disparities from the outset and should systematically track equity metrics over time to avoid perpetuating inequities in health care access.

## INTRODUCTION

The initial wave of coronavirus disease 2019 (COVID-19) redemonstrated and highlighted historical inequities in health by race, ethnicity, and other social indicators of vulnerability [1-3], prompting a range of efforts to design public health services that redress inequity in the COVID-19 response. Across a wide range of indicators, disease burden as measured by COVID-19 cases, hospitalizations, and mortality has disproportionately affected minoritized communities [1-3]. Initial responses to COVID-19 through established channels were thus accompanied by additional efforts to address the evolving disparities. Nevertheless, minoritized and vulnerable communities still had reduced access to testing and treatments and have endured disproportionate impacts of social distancing and lockdown policies on employment, education, and housing [4-8]. Against this backdrop, achieving equitable vaccinations has and continues to be one of the most critical public health challenges for mitigating the impact of the COVID-19 pandemic and achieving long-term control.

Closer examination of equity in the vaccine response evaluating the extent to which health systems performed in this domain is still necessary, and something that has not clearly documented in the literature to date. Whereas equality simply refers to provision of equal resources to every individual regardless of need, equitable approaches acknowledge that individuals will have different risks, needs, or opportunities and that access to or distribution of resources needs to take these differences into account. Strategies and frameworks to guide the equitable allocation and distributions of vaccine were developed for when vaccines for SARS CoV-2 became available in December 2020 [9,10], but empirical examination of how actual primary vaccine series and subsequent booster efforts aligned with the initial goals set out for equity are still needed. For example, several strategies proposed formally considering geography, social vulnerability, or race/ethnicity in plans to prioritize and distribute vaccinations in response to the known inequities in exposure risk and disease burden across these metrics [11-13]. Examinations of equity must thus document patterns of vaccinations across race/ethnicity, social vulnerability, geography, over time, and how they are delivered to understand the mechanisms that give rise to disparities and yield key insights to the success, failures, and steps for redress to achieve equitable vaccination strategies.

In this manuscript, we deepen our understanding of COVID-19 vaccine-related disparities by examining inequities in vaccination in the St. Louis and Kansas City regions in Missouri—regions with a history of health disparities—across several key metrics. We characterize rates of receiving the primary vaccine series and boosters over time, race/ethnicity, social vulnerability, disease burden, geography, and across vaccination location types. We use Lorenz curves and Gini coefficients—tools from economics commonly used to measure inequity in a population—to quantify and track inequities in COVID-19 vaccination over time relative to different metrics for conceptualizing equity [14]. The novel application of this methodology—which we previously used to characterize COVID-19 testing disparities [4]—has potential to yield deeper insights into the progress made towards vaccine equity in these regions that may then better inform health policy solutions to address remaining gaps.

## METHODS

### Ethics statement

The study was approved by the institutional review board at Washington University in St. Louis (IRB ID# 202009021). The research in this paper was not pre-specified and consists of secondary analysis of preexisting de-identified data. This manuscript was prepared according to STROBE guidelines (S1 STROBE Checklist).

### Study Setting and Data

We sought to assess disparities in COVID-19 vaccination across the 7 counties in the St. Louis region (St. Louis City, St. Louis County, St. Charles, Jefferson, Franklin, Lincoln, and Warren; total population 2,095,978: 19.2% Black, 73.1% White, 3.0% Hispanic, 3.2% Asian) and the 4 counties in the Kansas City region (Jackson, Clay, Cass, and Platte; total population 1,121,224: 16.8% Black, 73.2% White, 8.2% Hispanic, 2.0% Asian). These counties make up the broader metropolitan area located within Missouri for these two cities. Vaccines first became available on December 15, 2020 and all individuals became eligible on March 29, 2021. We used data from the Missouri State Department of Health and Senior Services on SARS-CoV-2 vaccines administered in Missouri to individuals 12 years old and up between December 15, 2020 and February 15, 2022. Reporting for vaccinations was mandated so this database is expected to contain near complete data on all vaccinations administered in Missouri. This individual-level dataset contains vaccination date, type, and dose number; administration site; and patient age, sex, race/ethnicity, and zip code, and was de-duplicated and cleaned by the Missouri State Department of Health and Senior Services. We used 2020 census data to obtain age-, sex-, and race-stratified zip code population estimates and 2018 American Community Surveys (ACS) data to obtain sociodemographic and socioeconomic characteristics of individual zip codes as well as the CDC’s social vulnerability index (SVI). The SVI is a composite metric that captures a community’s vulnerability to external stresses on human health and is calculated from 15 ACS variables measuring demographics, socioeconomic status, household composition, and infrastructure [15].

### Analyses

Our analyses seek to characterize patterns of disparities in receiving the primary vaccine series and boosters over time by examining rates of vaccination with respect to race/ethnicity and social vulnerability, changes in the type of locations vaccines were being administered, and the extent to which vaccine administration was equitable between zip codes. We adapted methods that we had previously used to assess disparities related to COVID-19 testing and extend them to COVID-19 vaccination [4].

First, we estimated the rates and cumulative incidence of COVID-19 vaccinations over time stratifying individuals by race/ethnicity (i.e., Black, White, Hispanic, or Asian) and whether they lived in zip codes with a low, medium, or high SVI (i.e., less than 0.333, 0.333 to 0.666, or greater than 0.666, respectively). We examined completion rates for the primary vaccine series (defined as 2 doses of either BNT162b2 mRNA [Pfizer] or mRNA-1273 [Moderna] or a single dose of Ad26.COV2.S [Johnson and Johnson]) and boosters (defined as a single dose of any vaccine after completing the primary series). Second, we examined the distribution by the type of sites at which individuals were receiving their primary vaccine series and boosters over time and by race/ethnicity and zip code-level SVI. We categorized vaccine administration sites into health facilities (e.g., clinics, hospitals, health system-affiliated sites) that administered either a small, medium, or large volume of vaccinations (i.e., less than 1000, 1000 to 10,000, or greater than 10,000 unique individuals vaccinated, respectively), public health departments (including mass vaccination sites), pharmacies, employer/school-based sites, and other (e.g., dialysis centers, home health, nursing homes, mental health/psychiatric facilities, and correctional facilities).

Third, we generated modified versions of Lorenz curves to assess the relative equity in the distribution of COVID-19 vaccinations across zip codes. Lorenz curves—originally developed by economists to graphically represent income equality—have more recently been leveraged as a tool for public health [14,16,17]. Lorenz curves are generated by plotting the cumulative proportion of the total population against the cumulative proportion of a resource after sorting values in ascending order. The curve follows a straight line at a 45-degree angle when a resource is equitably distributed across the population and becomes more convex with increasing inequity. In general, equitable vaccination strategies would seek to balance the number of vaccines with the overall risk of disease in a community, but the most appropriate metric of equity for so doing will depend on whether one considers the goal to be creating balance between vaccination rates relative to the total population, overall disease burden (i.e., number of COVID-19 cases or deaths), or risk factors (i.e., social vulnerability) in a community. To examine vaccine equity from these different perspectives, we adapted the Lorenz curve method to examine disparities in receiving the primary vaccine series and boosters relative to several relevant metrics: 1) the total population, 2) number of diagnosed COVID-19 cases, 3) number of COVID-19 deaths, and 4) population-level social vulnerability, which we defined as the zip code-level social vulnerability index multiplied by its population. For each curve, we calculated Gini coefficients—a measure of equality/inequality between 0 and 1, with 0 indicating perfect equality and 1 indicating perfect inequality—and assessed how these changed over time [18]. We also grouped zip codes into quartiles based on their position on Lorenz curves and assessed differences in zip code-level sociodemographic and socioeconomic characteristics using Kruskal-Wallis tests.

Fourth, we generated bubble plots to compare primary vaccine series and booster completion rates for Black, Hispanic, and Asian residents relative to White residents living in the same zip code. For these analyses, we only considered zip codes whose populations had at least 25 individuals for each of the race/ethnic groups being compared to avoid extreme outliers from small denominators.

Lastly, we performed univariate and multivariable mixed-effects Poisson regression to identify individual (e.g., sex, race/ethnicity, age) and zip-code level (e.g., SVI, racial makeup, health insurance coverage) factors independently associated with receiving the primary vaccine series and boosters; in multivariable models, we excluded zip code-level variables that would be expected to relate directly to SVI (e.g., poverty, median income). We applied an established method for using Poisson regression with robust variances to estimate risk ratios from binary outcomes [19,20]. We leveraged vaccination and 2020 census data to estimate the number of unvaccinated individuals across strata of age, sex, and race/ethnicity in each zip code. We visually assessed for linearity in the relationship between continuous variables and outcomes and presented variables with nonlinear relationships as categorical variables (i.e., age, zip code SVI). The effect of race/ethnicity and racism on health outcomes is mediated by (as opposed to confounded by) ecological structural factors such socioeconomic status; thus, unadjusted analyses assess the overall association with race/ethnicity and racism while adjusted analyses can be thought to assess the contribution of systemic racism that still remains even when adjusting away the mediating effects of measured ecological factors [21-23].

To account for missingness in race/ethnicity and patient zip code variables, we performed multiple imputation using multivariate normal imputation methods (n=50 imputations) [24-26]. For zip codes, we first transformed them to the latitude and longitude of their centroid, ran the multiple imputation model, and then transformed multiply imputed latitude and longitude values back into zip codes. Missingness was highly dependent on vaccination date and administration site, and thus the missing at random assumption required for unbiased imputation (i.e., that missingness was random conditional on all the variables included in the imputation model [administration site, vaccination date, sex, age, race/ethnicity, zip code latitude and longitude, type of vaccine]) was very plausible in our setting [24-26].

All analyses were conducted using Stata MP 17.0 and R 3.2.4.

## RESULTS

Between December 15, 2020 to February 15, 2022, 4,741,806 total COVID-19 vaccines were administered to 2,019,715 unique individuals across 7 counties in the St. Louis region and 4 counties in the Kansas City region. Among those receiving at least one dose in St. Louis and Kansas City, 1,762,508 (87.3%) completed the primary series and 871,896 (43.2%) had received a booster. Of those who completed the primary series, approximately 75% of individuals did so prior to June 15 and approximately 25% afterwards (Tables 1a and 1b, Table S1a and S1b).

**Table 1a.**
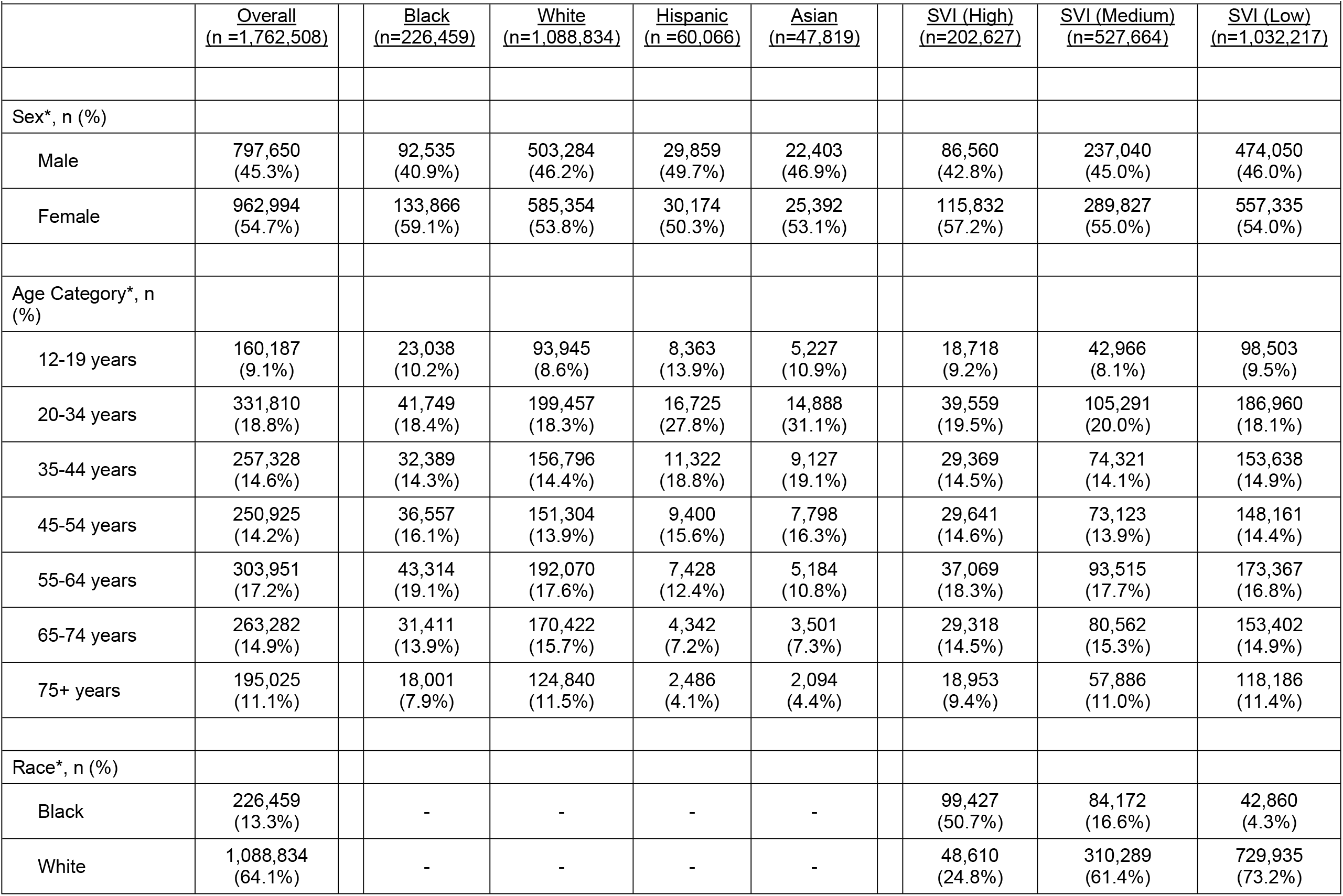
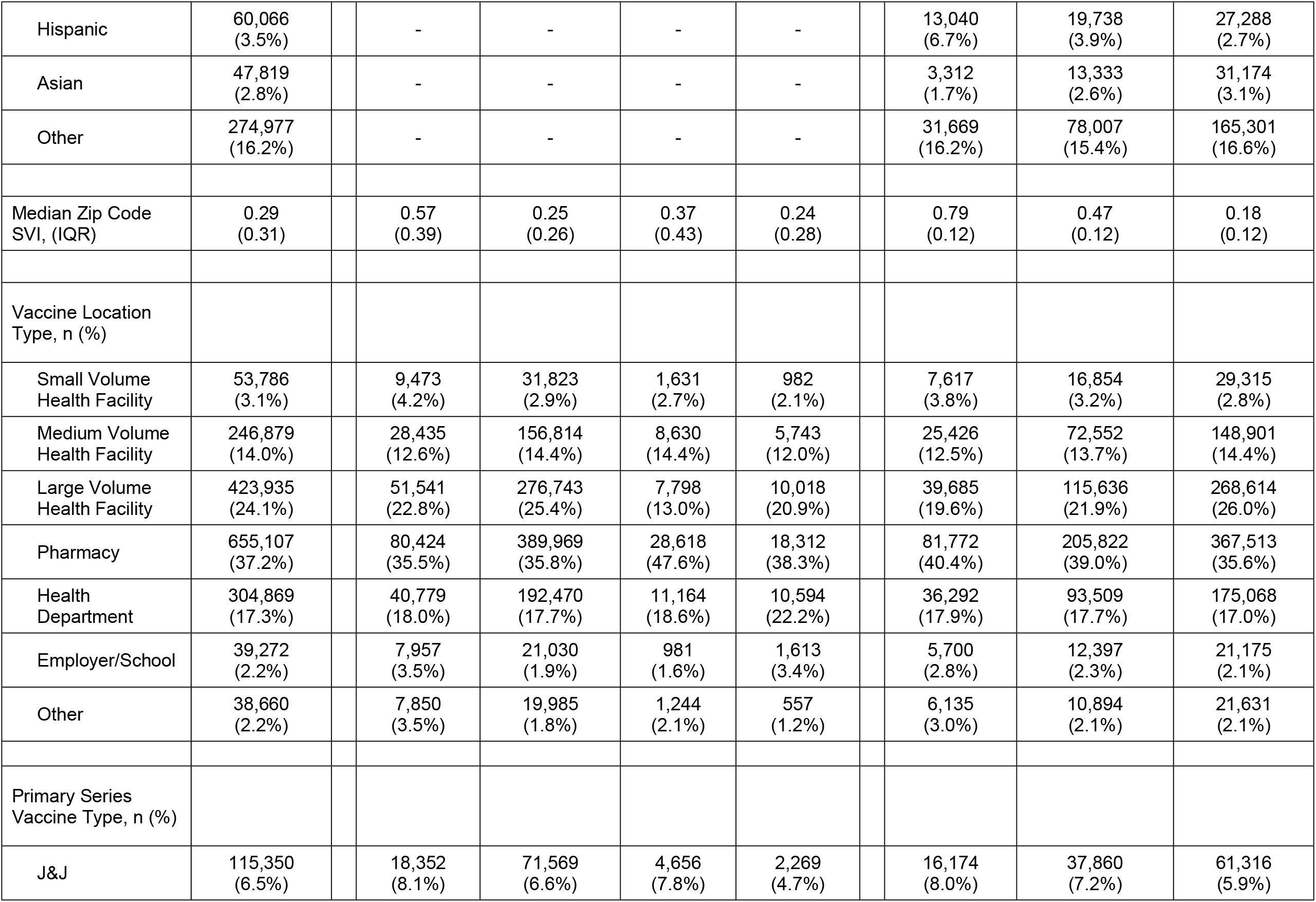
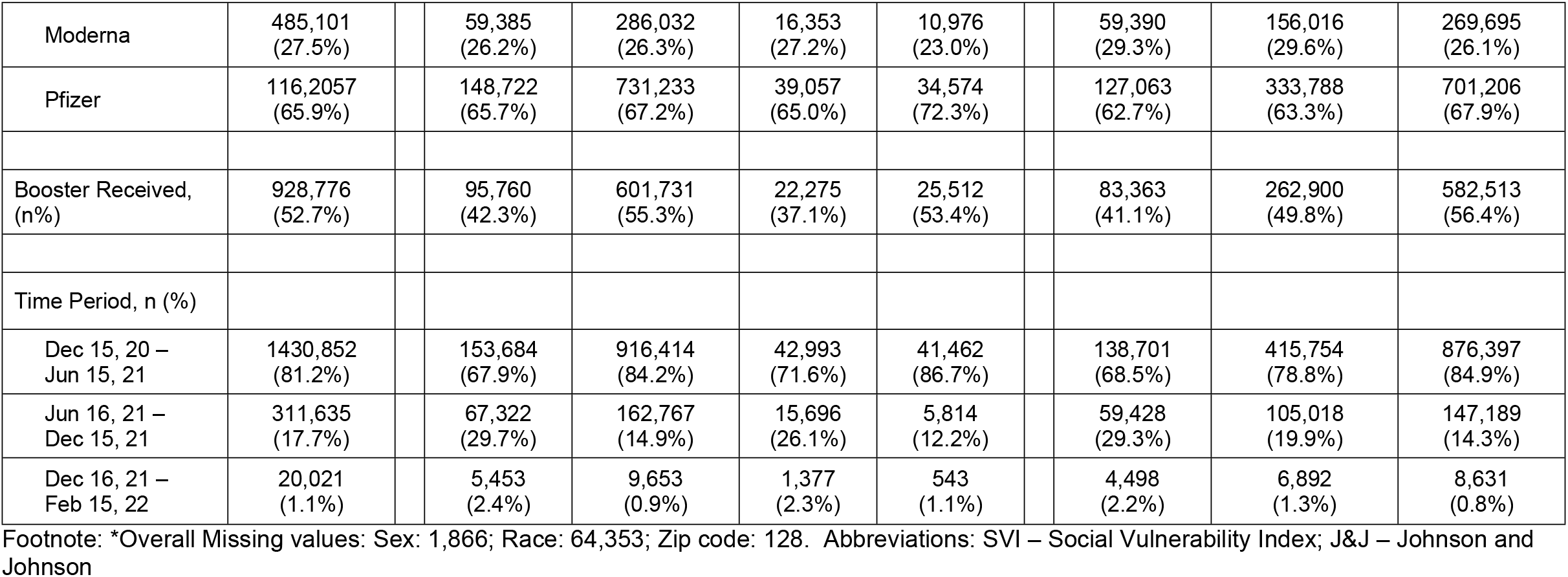
Characteristics of individuals completing primary series.

| Table 1a. Characteristics of individuals completing primary series |  |  |  |  |  |  |  |  |  |  |
| --- | --- | --- | --- | --- | --- | --- | --- | --- | --- | --- |
|  | Overall<br>(n =1,762,508) |  | Black<br>(n=226,459) | White<br>(n=1,088,834) | Hispanic<br>(n =60,066) | Asian<br>(n=47,819) |  | SVI (High)<br>(n=202,627) | SVI (Medium)<br>(n=527,664) | SVI (Low)<br>(n=1,032,217) |
| Sex*, n (%) |  |  |  |  |  |  |  |  |  |  |
| Male | 797,650<br>(45.3%) |  | 92,535<br>(40.9%) | 503,284<br>(46.2%) | 29,859<br>(49.7%) | 22,403<br>(46.9%) |  | 86,560<br>(42.8%) | 237,040<br>(45.0%) | 474,050<br>(46.0%) |
| Female | 962,994<br>(54.7%) |  | 133,866<br>(59.1%) | 585,354<br>(53.8%) | 30,174<br>(50.3%) | 25,392<br>(53.1%) |  | 115,832<br>(57.2%) | 289,827<br>(55.0%) | 557,335<br>(54.0%) |
| Age Category*, n (%) |  |  |  |  |  |  |  |  |  |  |
| 12-19 years | 160,187<br>(9.1%) |  | 23,038<br>(10.2%) | 93,945<br>(8.6%) | 8,363<br>(13.9%) | 5,227<br>(10.9%) |  | 18,718<br>(9.2%) | 42,966<br>(8.1%) | 98,503<br>(9.5%) |
| 20-34 years | 331,810<br>(18.8%) |  | 41,749<br>(18.4%) | 199,457<br>(18.3%) | 16,725<br>(27.8%) | 14,888<br>(31.1%) |  | 39,559<br>(19.5%) | 105,291<br>(20.0%) | 186,960<br>(18.1%) |
| 35-44 years | 257,328<br>(14.6%) |  | 32,389<br>(14.3%) | 156,796<br>(14.4%) | 11,322<br>(18.8%) | 9,127<br>(19.1%) |  | 29,369<br>(14.5%) | 74,321<br>(14.1%) | 153,638<br>(14.9%) |
| 45-54 years | 250,925<br>(14.2%) |  | 36,557<br>(16.1%) | 151,304<br>(13.9%) | 9,400<br>(15.6%) | 7,798<br>(16.3%) |  | 29,641<br>(14.6%) | 73,123<br>(13.9%) | 148,161<br>(14.4%) |
| 55-64 years | 303,951<br>(17.2%) |  | 43,314<br>(19.1%) | 192,070<br>(17.6%) | 7,428<br>(12.4%) | 5,184<br>(10.8%) |  | 37,069<br>(18.3%) | 93,515<br>(17.7%) | 173,367<br>(16.8%) |
| 65-74 years | 263,282<br>(14.9%) |  | 31,411<br>(13.9%) | 170,422<br>(15.7%) | 4,342<br>(7.2%) | 3,501<br>(7.3%) |  | 29,318<br>(14.5%) | 80,562<br>(15.3%) | 153,402<br>(14.9%) |
| 75+ years | 195,025<br>(11.1%) |  | 18,001<br>(7.9%) | 124,840<br>(11.5%) | 2,486<br>(4.1%) | 2,094<br>(4.4%) |  | 18,953<br>(9.4%) | 57,886<br>(11.0%) | 118,186<br>(11.4%) |
| Race*, n (%) |  |  |  |  |  |  |  |  |  |  |
| Black | 226,459<br>(13.3%) |  | - | - | - | - |  | 99,427<br>(50.7%) | 84,172<br>(16.6%) | 42,860<br>(4.3%) |
| White | 1,088,834<br>(64.1%) |  | - | - | - | - |  | 48,610<br>(24.8%) | 310,289<br>(61.4%) | 729,935<br>(73.2%) |

|  |  |  |  |  |  |  |  |  |
| --- | --- | --- | --- | --- | --- | --- | --- | --- |
| Hispanic | 60,066<br>(3.5%) | - | - | - | - | 13,040<br>(6.7%) | 19,738<br>(3.9%) | 27,288<br>(2.7%) |
| Asian | 47,819<br>(2.8%) | - | - | - | - | 3,312<br>(1.7%) | 13,333<br>(2.6%) | 31,174<br>(3.1%) |
| Other | 274,977<br>(16.2%) | - | - | - | - | 31,669<br>(16.2%) | 78,007<br>(15.4%) | 165,301<br>(16.6%) |
| Median Zip Code<br>SVI, (IQR) | 0.29<br>(0.31) | 0.57<br>(0.39) | 0.25<br>(0.26) | 0.37<br>(0.43) | 0.24<br>(0.28) | 0.79<br>(0.12) | 0.47<br>(0.12) | 0.18<br>(0.12) |
| Vaccine Location<br>Type, n (%) |  |  |  |  |  |  |  |  |
| Small Volume<br>Health Facility | 53,786<br>(3.1%) | 9,473<br>(4.2%) | 31,823<br>(2.9%) | 1,631<br>(2.7%) | 982<br>(2.1%) | 7,617<br>(3.8%) | 16,854<br>(3.2%) | 29,315<br>(2.8%) |
| Medium Volume<br>Health Facility | 246,879<br>(14.0%) | 28,435<br>(12.6%) | 156,814<br>(14.4%) | 8,630<br>(14.4%) | 5,743<br>(12.0%) | 25,426<br>(12.5%) | 72,552<br>(13.7%) | 148,901<br>(14.4%) |
| Large Volume<br>Health Facility | 423,935<br>(24.1%) | 51,541<br>(22.8%) | 276,743<br>(25.4%) | 7,798<br>(13.0%) | 10,018<br>(20.9%) | 39,685<br>(19.6%) | 115,636<br>(21.9%) | 268,614<br>(26.0%) |
| Pharmacy | 655,107<br>(37.2%) | 80,424<br>(35.5%) | 389,969<br>(35.8%) | 28,618<br>(47.6%) | 18,312<br>(38.3%) | 81,772<br>(40.4%) | 205,822<br>(39.0%) | 367,513<br>(35.6%) |
| Health<br>Department | 304,869<br>(17.3%) | 40,779<br>(18.0%) | 192,470<br>(17.7%) | 11,164<br>(18.6%) | 10,594<br>(22.2%) | 36,292<br>(17.9%) | 93,509<br>(17.7%) | 175,068<br>(17.0%) |
| Employer/School | 39,272<br>(2.2%) | 7,957<br>(3.5%) | 21,030<br>(1.9%) | 981<br>(1.6%) | 1,613<br>(3.4%) | 5,700<br>(2.8%) | 12,397<br>(2.3%) | 21,175<br>(2.1%) |
| Other | 38,660<br>(2.2%) | 7,850<br>(3.5%) | 19,985<br>(1.8%) | 1,244<br>(2.1%) | 557<br>(1.2%) | 6,135<br>(3.0%) | 10,894<br>(2.1%) | 21,631<br>(2.1%) |
| Primary Series<br>Vaccine Type, n (%) |  |  |  |  |  |  |  |  |
| J&J | 115,350<br>(6.5%) | 18,352<br>(8.1%) | 71,569<br>(6.6%) | 4,656<br>(7.8%) | 2,269<br>(4.7%) | 16,174<br>(8.0%) | 37,860<br>(7.2%) | 61,316<br>(5.9%) |
| Moderna | 485,101<br>(27.5%) |  | 59,385<br>(26.2%) | 286,032<br>(26.3%) | 16,353<br>(27.2%) | 10,976<br>(23.0%) |  | 59,390<br>(29.3%) | 156,016<br>(29.6%) | 269,695<br>(26.1%) |
| Pfizer | 116,2057<br>(65.9%) |  | 148,722<br>(65.7%) | 731,233<br>(67.2%) | 39,057<br>(65.0%) | 34,574<br>(72.3%) |  | 127,063<br>(62.7%) | 333,788<br>(63.3%) | 701,206<br>(67.9%) |
| Booster Received,<br>(n%) | 928,776<br>(52.7%) |  | 95,760<br>(42.3%) | 601,731<br>(55.3%) | 22,275<br>(37.1%) | 25,512<br>(53.4%) |  | 83,363<br>(41.1%) | 262,900<br>(49.8%) | 582,513<br>(56.4%) |
| Time Period, n (%) |  |  |  |  |  |  |  |  |  |  |
| Dec 15, 20 –<br>Jun 15, 21 | 1430,852<br>(81.2%) |  | 153,684<br>(67.9%) | 916,414<br>(84.2%) | 42,993<br>(71.6%) | 41,462<br>(86.7%) |  | 138,701<br>(68.5%) | 415,754<br>(78.8%) | 876,397<br>(84.9%) |
| Jun 16, 21 –<br>Dec 15, 21 | 311,635<br>(17.7%) |  | 67,322<br>(29.7%) | 162,767<br>(14.9%) | 15,696<br>(26.1%) | 5,814<br>(12.2%) |  | 59,428<br>(29.3%) | 105,018<br>(19.9%) | 147,189<br>(14.3%) |
| Dec 16, 21 –<br>Feb 15, 22 | 20,021<br>(1.1%) |  | 5,453<br>(2.4%) | 9,653<br>(0.9%) | 1,377<br>(2.3%) | 543<br>(1.1%) |  | 4,498<br>(2.2%) | 6,892<br>(1.3%) | 8,631<br>(0.8%) |

Footnote: \*Overall Missing values: Sex: 1,866; Race: 64,353; Zip code: 128. Abbreviations: SVI – Social Vulnerability Index; J&J – Johnson and Johnson

**Table 1b.**
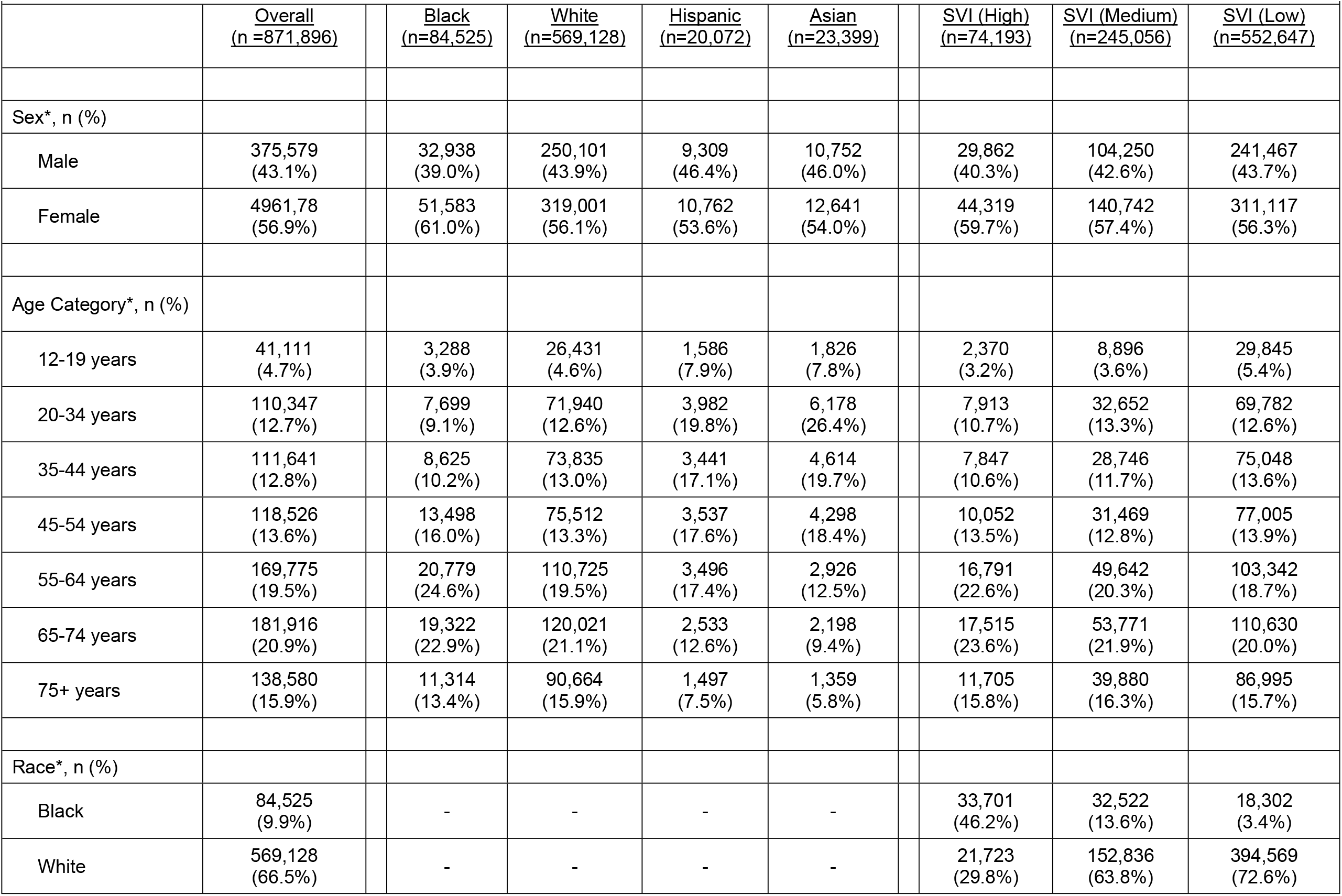
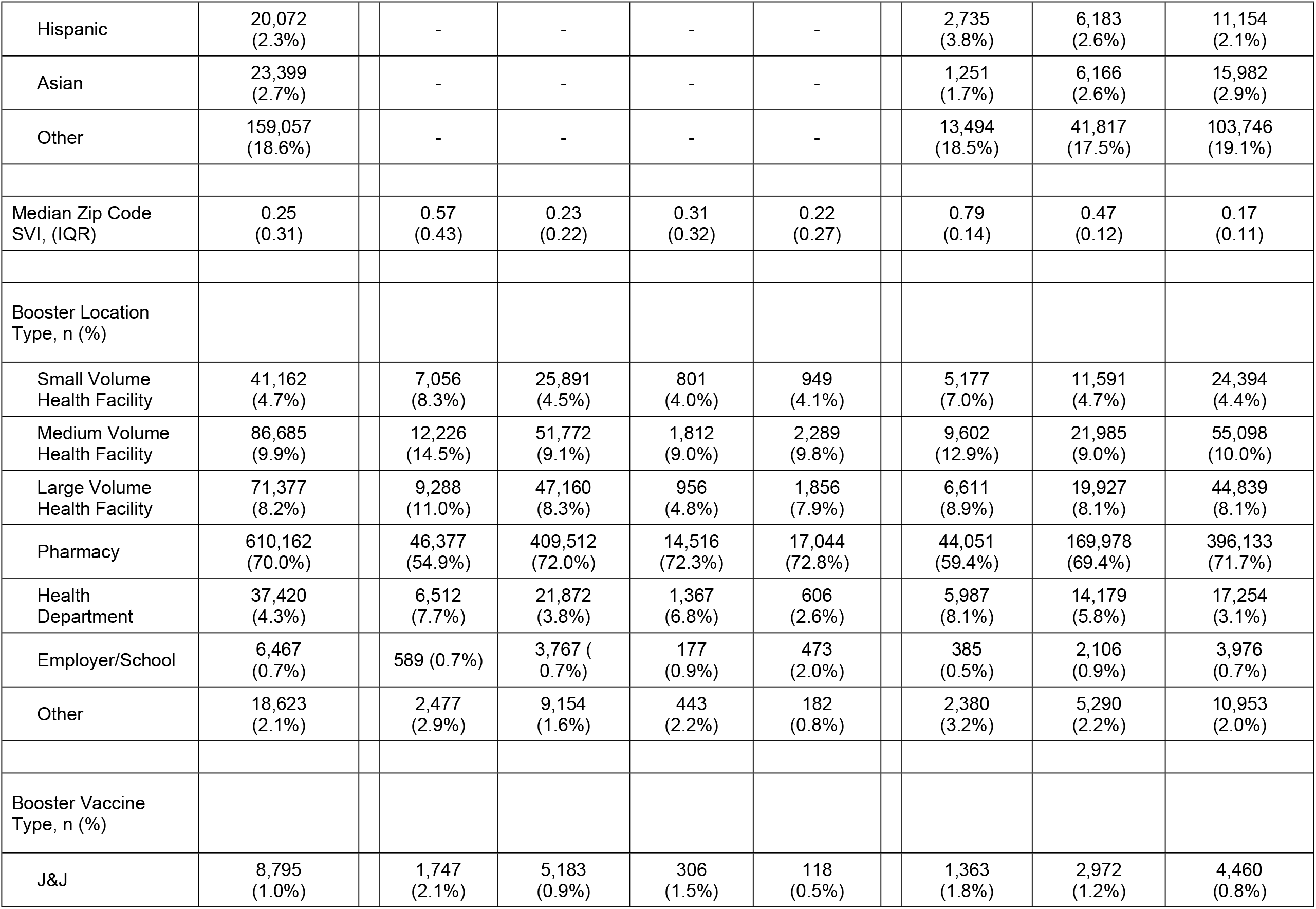
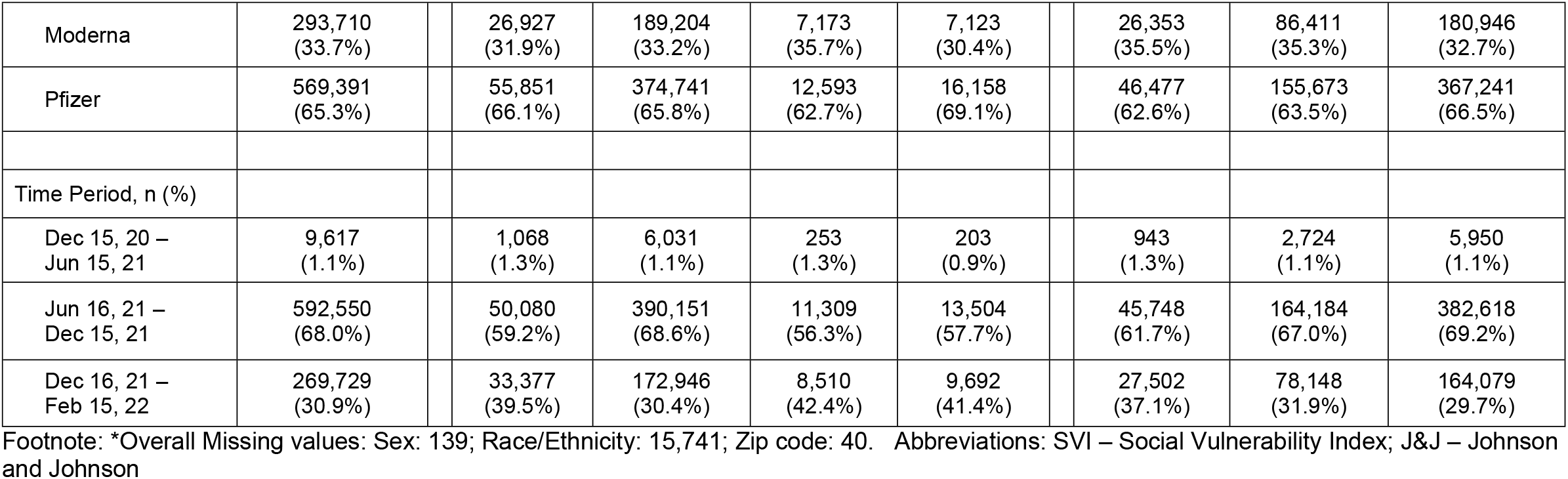
Characteristics of individuals receiving a booster vaccination.

|  | <u>Overall</u><br>(n =871,896) | <u>Black</u><br>(n=84,525) | <u>White</u><br>(n=569,128) | <u>Hispanic</u><br>(n=20,072) | <u>Asian</u><br>(n=23,399) | <u>SVI (High)</u><br>(n=74,193) | <u>SVI (Medium)</u><br>(n=245,056) | <u>SVI (Low)</u><br>(n=552,647) |
| --- | --- | --- | --- | --- | --- | --- | --- | --- |
| Sex*, n (%) |  |  |  |  |  |  |  |  |
| Male | 375,579<br>(43.1%) | 32,938<br>(39.0%) | 250,101<br>(43.9%) | 9,309<br>(46.4%) | 10,752<br>(46.0%) | 29,862<br>(40.3%) | 104,250<br>(42.6%) | 241,467<br>(43.7%) |
| Female | 4961,78<br>(56.9%) | 51,583<br>(61.0%) | 319,001<br>(56.1%) | 10,762<br>(53.6%) | 12,641<br>(54.0%) | 44,319<br>(59.7%) | 140,742<br>(57.4%) | 311,117<br>(56.3%) |
| Age Category*, n (%) |  |  |  |  |  |  |  |  |
| 12-19 years | 41,111<br>(4.7%) | 3,288<br>(3.9%) | 26,431<br>(4.6%) | 1,586<br>(7.9%) | 1,826<br>(7.8%) | 2,370<br>(3.2%) | 8,896<br>(3.6%) | 29,845<br>(5.4%) |
| 20-34 years | 110,347<br>(12.7%) | 7,699<br>(9.1%) | 71,940<br>(12.6%) | 3,982<br>(19.8%) | 6,178<br>(26.4%) | 7,913<br>(10.7%) | 32,652<br>(13.3%) | 69,782<br>(12.6%) |
| 35-44 years | 111,641<br>(12.8%) | 8,625<br>(10.2%) | 73,835<br>(13.0%) | 3,441<br>(17.1%) | 4,614<br>(19.7%) | 7,847<br>(10.6%) | 28,746<br>(11.7%) | 75,048<br>(13.6%) |
| 45-54 years | 118,526<br>(13.6%) | 13,498<br>(16.0%) | 75,512<br>(13.3%) | 3,537<br>(17.6%) | 4,298<br>(18.4%) | 10,052<br>(13.5%) | 31,469<br>(12.8%) | 77,005<br>(13.9%) |
| 55-64 years | 169,775<br>(19.5%) | 20,779<br>(24.6%) | 110,725<br>(19.5%) | 3,496<br>(17.4%) | 2,926<br>(12.5%) | 16,791<br>(22.6%) | 49,642<br>(20.3%) | 103,342<br>(18.7%) |
| 65-74 years | 181,916<br>(20.9%) | 19,322<br>(22.9%) | 120,021<br>(21.1%) | 2,533<br>(12.6%) | 2,198<br>(9.4%) | 17,515<br>(23.6%) | 53,771<br>(21.9%) | 110,630<br>(20.0%) |
| 75+ years | 138,580<br>(15.9%) | 11,314<br>(13.4%) | 90,664<br>(15.9%) | 1,497<br>(7.5%) | 1,359<br>(5.8%) | 11,705<br>(15.8%) | 39,880<br>(16.3%) | 86,995<br>(15.7%) |
| Race*, n (%) |  |  |  |  |  |  |  |  |
| Black | 84,525<br>(9.9%) | - | - | - | - | 33,701<br>(46.2%) | 32,522<br>(13.6%) | 18,302<br>(3.4%) |
| White | 569,128<br>(66.5%) | - | - | - | - | 21,723<br>(29.8%) | 152,836<br>(63.8%) | 394,569<br>(72.6%) |
| Hispanic | 20,072<br>(2.3%) | - | - | - | - | 2,735<br>(3.8%) | 6,183<br>(2.6%) | 11,154<br>(2.1%) |
| Asian | 23,399<br>(2.7%) | - | - | - | - | 1,251<br>(1.7%) | 6,166<br>(2.6%) | 15,982<br>(2.9%) |
| Other | 159,057<br>(18.6%) | - | - | - | - | 13,494<br>(18.5%) | 41,817<br>(17.5%) | 103,746<br>(19.1%) |
| Median Zip Code<br>SVI, (IQR) | 0.25<br>(0.31) | 0.57<br>(0.43) | 0.23<br>(0.22) | 0.31<br>(0.32) | 0.22<br>(0.27) | 0.79<br>(0.14) | 0.47<br>(0.12) | 0.17<br>(0.11) |
| Booster Location<br>Type, n (%) |  |  |  |  |  |  |  |  |
| Small Volume<br>Health Facility | 41,162<br>(4.7%) | 7,056<br>(8.3%) | 25,891<br>(4.5%) | 801<br>(4.0%) | 949<br>(4.1%) | 5,177<br>(7.0%) | 11,591<br>(4.7%) | 24,394<br>(4.4%) |
| Medium Volume<br>Health Facility | 86,685<br>(9.9%) | 12,226<br>(14.5%) | 51,772<br>(9.1%) | 1,812<br>(9.0%) | 2,289<br>(9.8%) | 9,602<br>(12.9%) | 21,985<br>(9.0%) | 55,098<br>(10.0%) |
| Large Volume<br>Health Facility | 71,377<br>(8.2%) | 9,288<br>(11.0%) | 47,160<br>(8.3%) | 956<br>(4.8%) | 1,856<br>(7.9%) | 6,611<br>(8.9%) | 19,927<br>(8.1%) | 44,839<br>(8.1%) |
| Pharmacy | 610,162<br>(70.0%) | 46,377<br>(54.9%) | 409,512<br>(72.0%) | 14,516<br>(72.3%) | 17,044<br>(72.8%) | 44,051<br>(59.4%) | 169,978<br>(69.4%) | 396,133<br>(71.7%) |
| Health<br>Department | 37,420<br>(4.3%) | 6,512<br>(7.7%) | 21,872<br>(3.8%) | 1,367<br>(6.8%) | 606<br>(2.6%) | 5,987<br>(8.1%) | 14,179<br>(5.8%) | 17,254<br>(3.1%) |
| Employer/School | 6,467<br>(0.7%) | 589 (0.7%) | 3,767 (0.7%) | 177<br>(0.9%) | 473<br>(2.0%) | 385<br>(0.5%) | 2,106<br>(0.9%) | 3,976<br>(0.7%) |
| Other | 18,623<br>(2.1%) | 2,477<br>(2.9%) | 9,154<br>(1.6%) | 443<br>(2.2%) | 182<br>(0.8%) | 2,380<br>(3.2%) | 5,290<br>(2.2%) | 10,953<br>(2.0%) |
| Booster Vaccine<br>Type, n (%) |  |  |  |  |  |  |  |  |
| J&J | 8,795<br>(1.0%) | 1,747<br>(2.1%) | 5,183<br>(0.9%) | 306<br>(1.5%) | 118<br>(0.5%) | 1,363<br>(1.8%) | 2,972<br>(1.2%) | 4,460<br>(0.8%) |

|  |  |  |  |  |  |  |  |  |  |  |
| --- | --- | --- | --- | --- | --- | --- | --- | --- | --- | --- |
| Moderna | 293,710<br>(33.7%) |  | 26,927<br>(31.9%) | 189,204<br>(33.2%) | 7,173<br>(35.7%) | 7,123<br>(30.4%) |  | 26,353<br>(35.5%) | 86,411<br>(35.3%) | 180,946<br>(32.7%) |
| Pfizer | 569,391<br>(65.3%) |  | 55,851<br>(66.1%) | 374,741<br>(65.8%) | 12,593<br>(62.7%) | 16,158<br>(69.1%) |  | 46,477<br>(62.6%) | 155,673<br>(63.5%) | 367,241<br>(66.5%) |
| Time Period, n (%) |  |  |  |  |  |  |  |  |  |  |
| Dec 15, 20 –<br>Jun 15, 21 | 9,617<br>(1.1%) |  | 1,068<br>(1.3%) | 6,031<br>(1.1%) | 253<br>(1.3%) | 203<br>(0.9%) |  | 943<br>(1.3%) | 2,724<br>(1.1%) | 5,950<br>(1.1%) |
| Jun 16, 21 –<br>Dec 15, 21 | 592,550<br>(68.0%) |  | 50,080<br>(59.2%) | 390,151<br>(68.6%) | 11,309<br>(56.3%) | 13,504<br>(57.7%) |  | 45,748<br>(61.7%) | 164,184<br>(67.0%) | 382,618<br>(69.2%) |
| Dec 16, 21 –<br>Feb 15, 22 | 269,729<br>(30.9%) |  | 33,377<br>(39.5%) | 172,946<br>(30.4%) | 8,510<br>(42.4%) | 9,692<br>(41.4%) |  | 27,502<br>(37.1%) | 78,148<br>(31.9%) | 164,079<br>(29.7%) |

Footnote: \*Overall Missing values: Sex: 139; Race/Ethnicity: 15,741; Zip code: 40. Abbreviations: SVI – Social Vulnerability Index; J&J – Johnson and Johnson

### Rates of COVID-19 Primary and Booster Vaccinations by Race/Ethnicity and SVI over Time

The rate of primary COVID-19 vaccinations steadily increased until peaking in mid-April. This was followed by rapid decline with smaller upticks at the end of May and then during the Delta wave beginning in July; there was no corresponding uptick in vaccination rates during the Omicron wave beginning in mid-December (Figure 1, Table/Figures S2-S5). Up through April, White individuals from zip codes with low SVIs were vaccinated at a rate greater than 2 times that of Black and Hispanic individuals from high SVI zip codes, but the rate ratio declined over time. Asian individuals from all zip codes were vaccinated at the highest rates. During the same early period, Black and Hispanic individuals from low-SVI zip codes were vaccinated at rates somewhat similar to or higher than White individuals from medium and high SVI zip codes. After June, Black and Hispanic individuals from high, medium, and low SVI zip codes were vaccinated at higher rates than White individuals, although this was also during periods with lower absolute numbers of vaccinations (Figure 1, Table/Figures S2-S5). Patterns were largely similar across St. Louis and Kansas City (Figures S4a-S4b).

**Figure 1:**
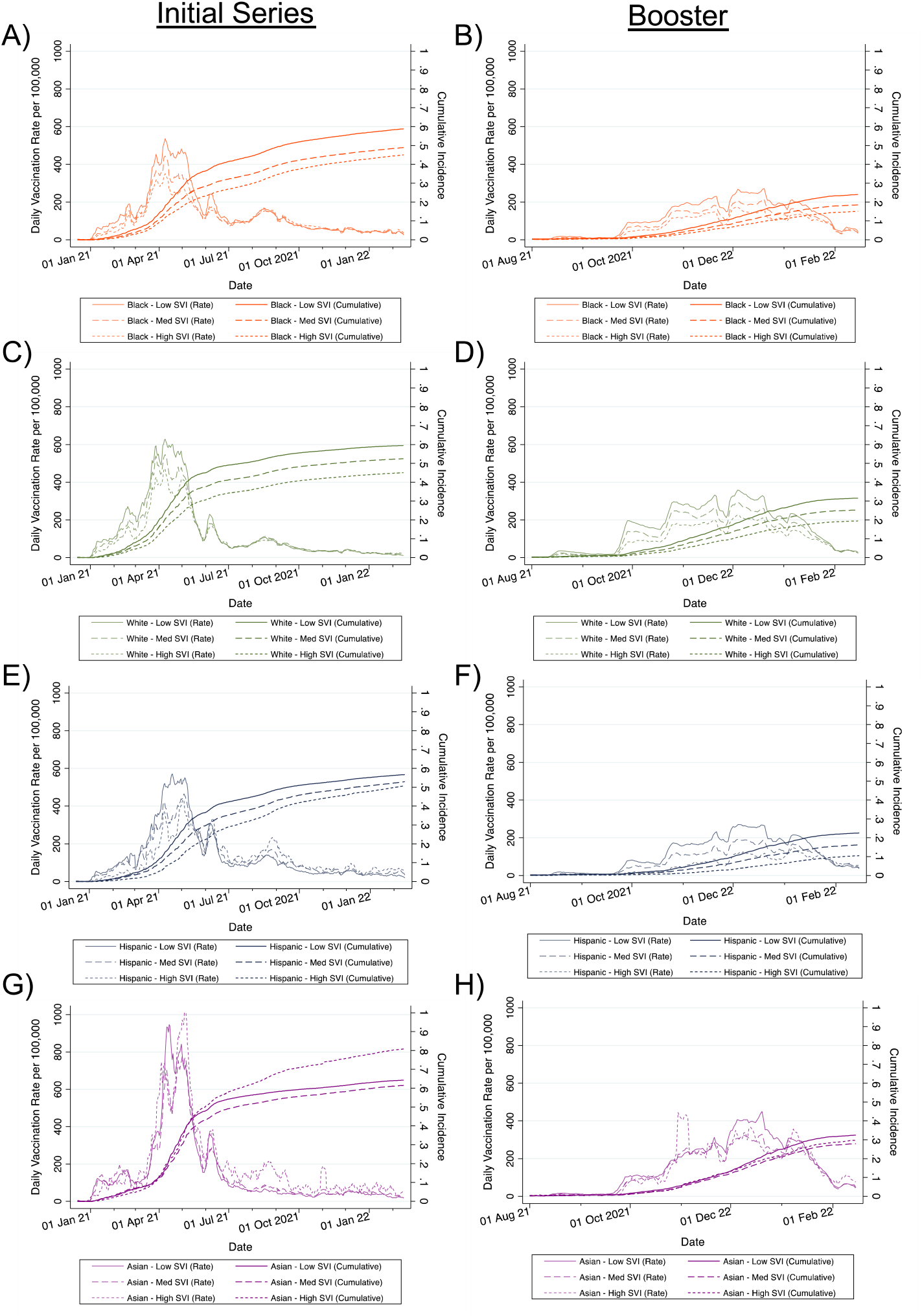
Rates and Cumulative Incidence of Receiving the Primary COVID-19 Vaccination Series and Boosters by Race/Ethnicity and SVI over Time. Estimates represent 7-day moving averages derived from multiply imputed datasets. Denominators represent the total population greater than or equal to 12 years old. Low SVI indicates zip codes with SVIs less than 0.333, medium SVI indicates SVIs between 0.333 and 0.666, and high SVI indicates SVIs greater than or equal to 0.666. SVI=Social Vulnerability Index.

Booster rates increased starting in October 2021 and peaked in early December at the beginning of the Omicron wave, albeit at much lower levels than for the primary vaccine series, and started to decline in January 2022. Patterns of disparities across race/ethnicity were similar for boosters compared to completion of the primary series (Figure 1, Table/Figures S2-S5).

### Locations of COVID-19 Vaccinations Over Time

Early during the vaccination campaign, the vast majority of vaccines were delivered through medium and large volume health facilities (Figure 2). From February through April, a substantial proportion were also delivered through public health departments (including mass vaccination sites). After April, the proportion of vaccines being administered through pharmacies steadily increased accounting for about 70% of vaccines administered after July. Black individuals received comparatively more vaccines through employer/school sponsored sites, small volume health facilities, or other facilities such as dialysis centers, home health, and nursing homes and fewer from pharmacies and health departments. Hispanic and Asian individuals received comparatively more vaccines through pharmacies and health departments; Hispanic individuals also received relatively few vaccines from large volume health facilities. Again, patterns were qualitatively similar for boosters (Figure 2).

**Figure 2:**
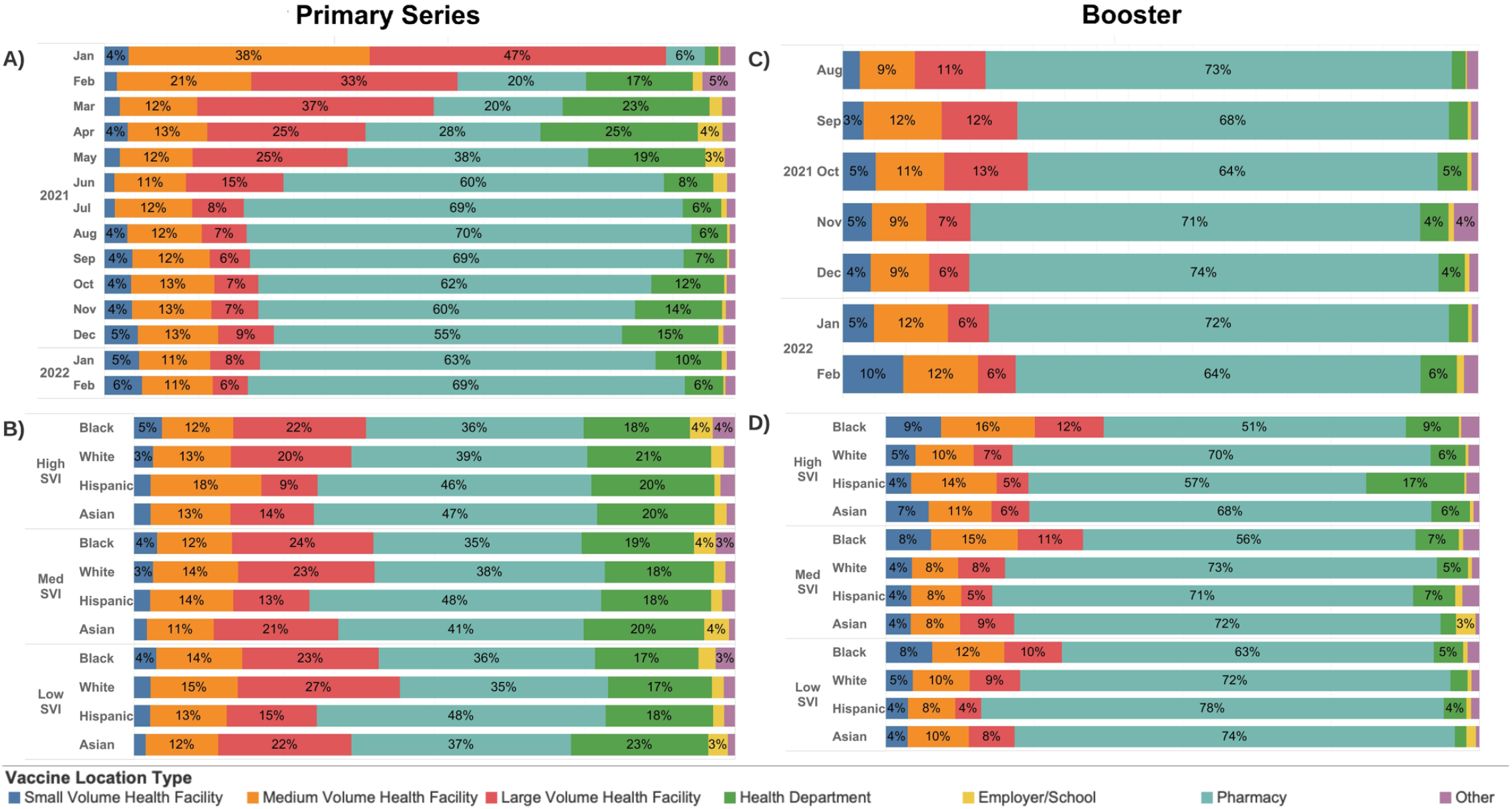
Distribution of Primary COVID-19 Vaccine Series and Boosters by Location Type over time, SVI, and race/ethnicity. Low SVI indicates zip codes with SVIs less than 0.333, medium SVI is between 0.333 and 0.666, and high SVI is greater than or equal to 0.666. Health facilities were categorized as small-, medium-, and large-volume based on whether they vaccinated less than 1000, 1000 to 10,000, or greater than 10,000 unique individuals. Other facilities included dialysis centers, home health, nursing homes, mental health/psychiatric facilities, and correctional facilities. Primary series vaccines were allocated to the location where the series was completed. SVI=Social Vulnerability Index.

### COVID-19 Vaccine Disparities across Zip Codes using Lorenz Curves

Modified Lorenz curves depict the distribution of COVID-19 vaccinations with respect to the total population, diagnosed COVID-19 cases, deaths, and population-level SVI across zip codes (Figure 3). For the primary vaccine series, zip codes in the quartile with the lowest rates of vaccinations accounted for 19.3%, 18.1%, 10.8%, and 8.8% of vaccines but represented 25% of either the total population, cases, deaths, or population-level SVI, respectively. These zip codes, in general, had higher proportions of Black residents, lower median incomes, higher rates of poverty, lower rates of health insurance coverage, a higher proportion of residents employed in the service sector, and higher COVID-19 deaths (Figure 3, Tables S6-S9). In contrast, zip codes with the highest rates of vaccinations accounted for 30.7%, 35.0%, 44.2%, and 56.1% of vaccinations, but represented 25% of either the total population, cases, deaths, or population-SVI, respectively. These zip codes tended to have a lower percentage of Black residents and be more socioeconomically advantaged (Figure 3, Tables S6-S9). These patterns were similar, but demonstrated a greater magnitude of disparities for boosters (Figure 3, Tables S6-S9).

**Figure 3:**
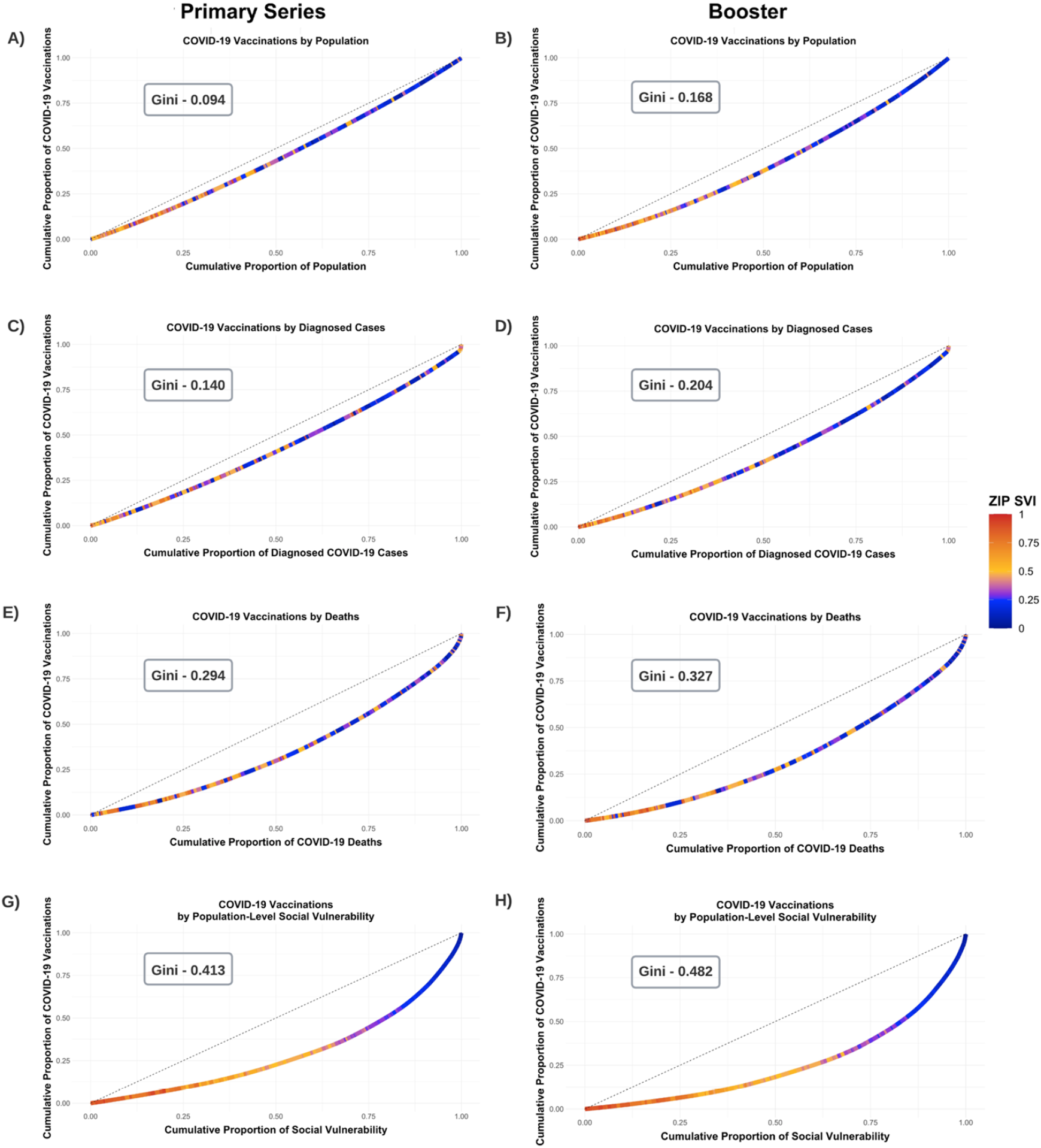
Lorenz Curves of Disparities in COVID-19 Vaccinations. This figure depicts modified Lorenz curve examining disparities in COVID-19 vaccinations as of February 15, 2022. The units of analysis are zip codes and they are color-coded by their SVI. The dashed line represents equitable distribution where 50% of vaccinations would be conducting in zip codes accounting for either 50% of the population, cases, deaths, or total social vulnerability. Lorenz curves measure disparities in the distribution of receiving 1) the primary vaccine series and 2) a booster relative to the total population above 12 years old (Panels A, B), diagnosed COVID-19 cases (Panels C, D), deaths, (Panels E, F), and total social vulnerability (Panels G, H).

When examining changes in Gini coefficients and vaccine inequities between zip codes over time, inequities were extremely high during the initial periods of the primary series rollout, but began to slowly improve after February 2021 relative to population, deaths, and total social vulnerability, but improvements relative to diagnosed cases plateaued around May 2021. Nevertheless, these improvements were slow and modest and vaccine inequities between zip codes remained substantial all metrics through to January 2022 (Figures 4 and S10). With respect to boosters, Gini coefficients once again were very high in the beginning of rollout, followed by slow improvement with relative to population, cases, and deaths; Ginis did not improve (and even worsened initially) relative to total social vulnerability (Figures 4 and S11). There were limited improvements after December 2022 during the Omicron wave.

**Figure 4:**
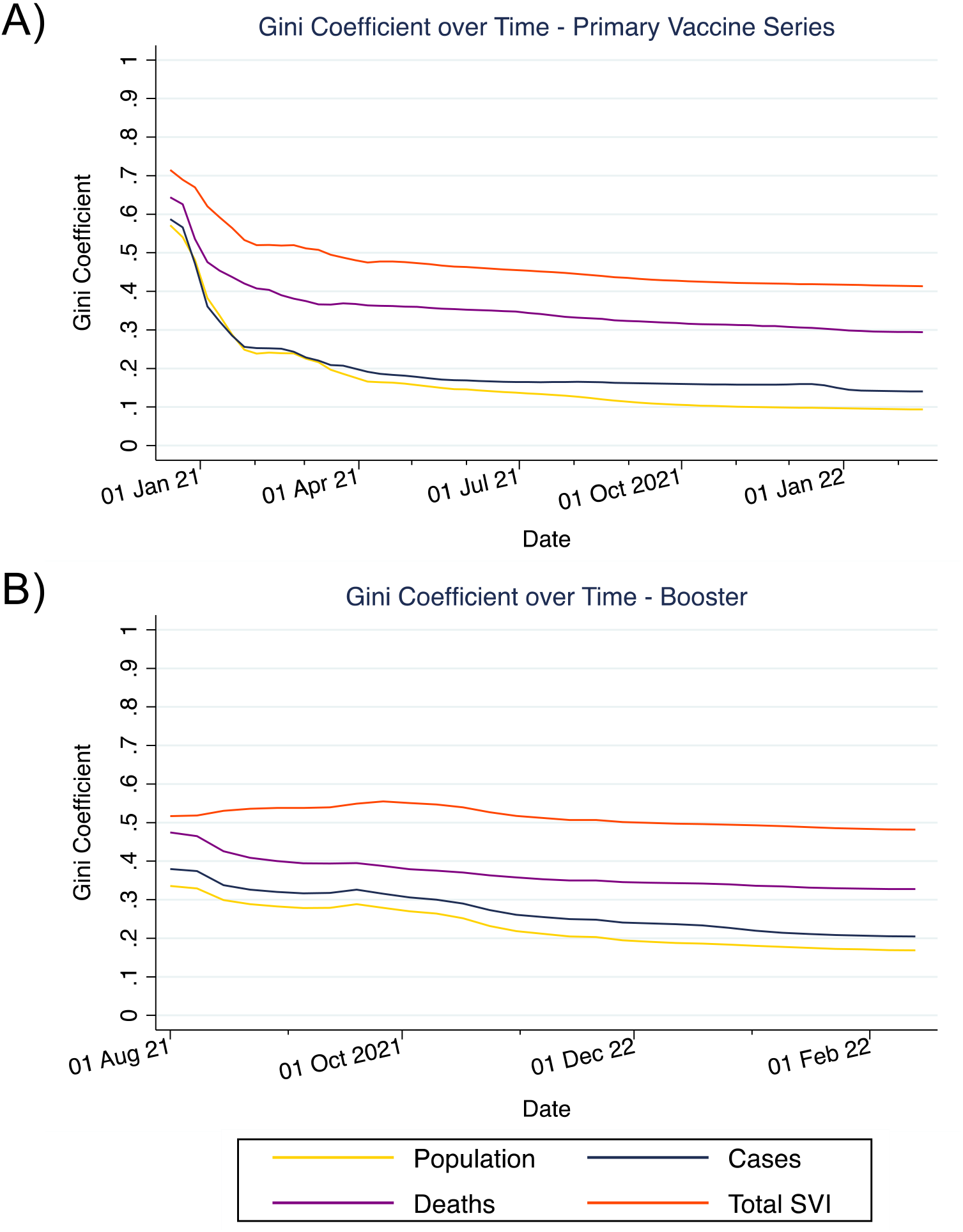
Temporal Trends in COVID-19 Vaccine Inequities. This figure depicts trends in the Gini coefficients over time for inequities in receiving 1) the primary vaccine series (Panel A) and 2) a booster (Panel B) relative to population, diagnosed COVID-19 cases, COVID-19 deaths, and population-level social vulnerability. Gini coefficients were calculated on a weekly basis from Lorenz curves generated up through that time interval.

### COVID-19 Vaccine Disparities within Zip Codes

Black, Hispanic, and Asian individuals generally had lower rates of primary series completion compared to White individuals residing in the same zip codes in zip codes with lower vaccination coverage (which also tended to have higher SVIs) (Figures 5 and S12). However, in zip codes with high vaccine coverage (which also tended to have low SVIs), Black, Hispanic, and Asian communities often had higher primary series completion compare to White communities in the same zip code. For boosters, Black and Hispanic communities had lower vaccination rates compared to White communities across most zip codes, although Asian communities trended slightly towards have higher booster rates (Figures 5 and S12).

**Figure 5:**
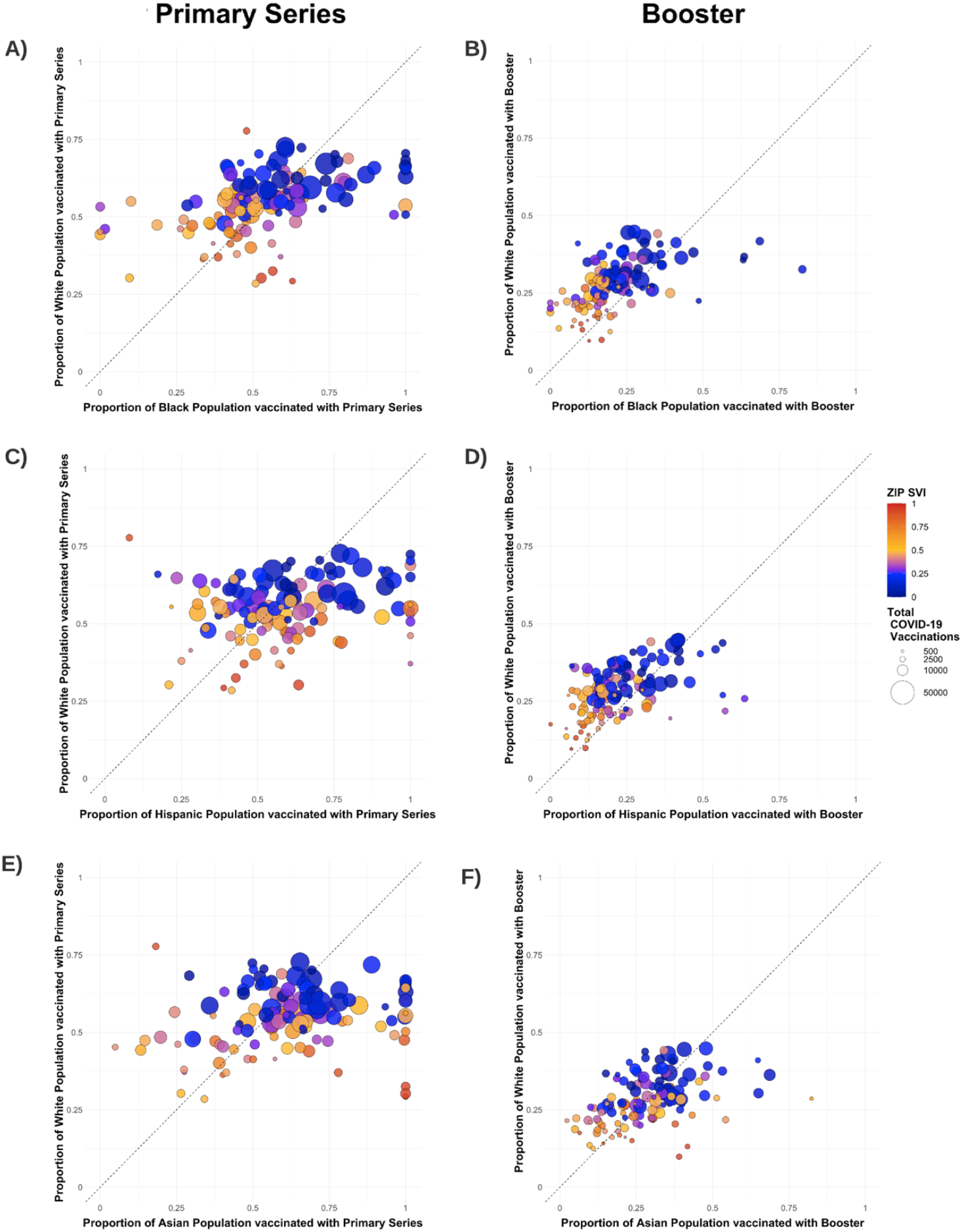
Disparities in COVID-19 Primary Vaccine Series and Boosters Among Black, Hispanic, and Asian versus White residents of the same zip code. This figure depicts vaccination rates for the primary series and boosters for Black (Panel A,B), Hispanic (C, D), and Asian (E,F) residents compared to the White residents of the same zip code. Each marker represents a single zip code. Markers are color-coded by the zip code SVI and sized by the total number of vaccines administered in the zip code. The dashed line represents equitable vaccine distribution between racial/communities being compared. Zip codes falling above the dashed line indicates that there was decreased vaccination in Black, Hispanic, or Asian residents as opposed to White residents (and vice versa).

### Factors Associated with Receiving the Primary Vaccine Series and Boosters

In multivariable mixed-effects Poisson regression, Black and Hispanic individuals had slightly lower rates of completing the primary vaccine series compared to White individuals (aRRs 0.94 [95% CI 0.93-0.94] and 0.96 [95% CI 0.95-0.97], respectively), while Asian individuals had slightly higher rates (aRRs 1.03 [95% CI 1.02-1.03]). Living in medium and high SVI zip codes was also associated with lower vaccination rates compared to low SVI zip codes (aRRs 0.92 [95% CI 0.91-0.92] and 0.88 [95% CI 0.88-0.89]) (Table 2). Additional factors associated with increased vaccination were being female, being over 55, or between 12 to 19 years old (as compared to 45 to 55 years old); 20 to 34 years olds had decreased vaccination rates. Differences in receipt of a booster vaccine were substantially higher across race, age, sex, and zip code SVI compared to the differences in completion of the primary vaccine series, except that 12- to 19-year-olds were less likely to receive a booster (Table 2).

**Table 2.**
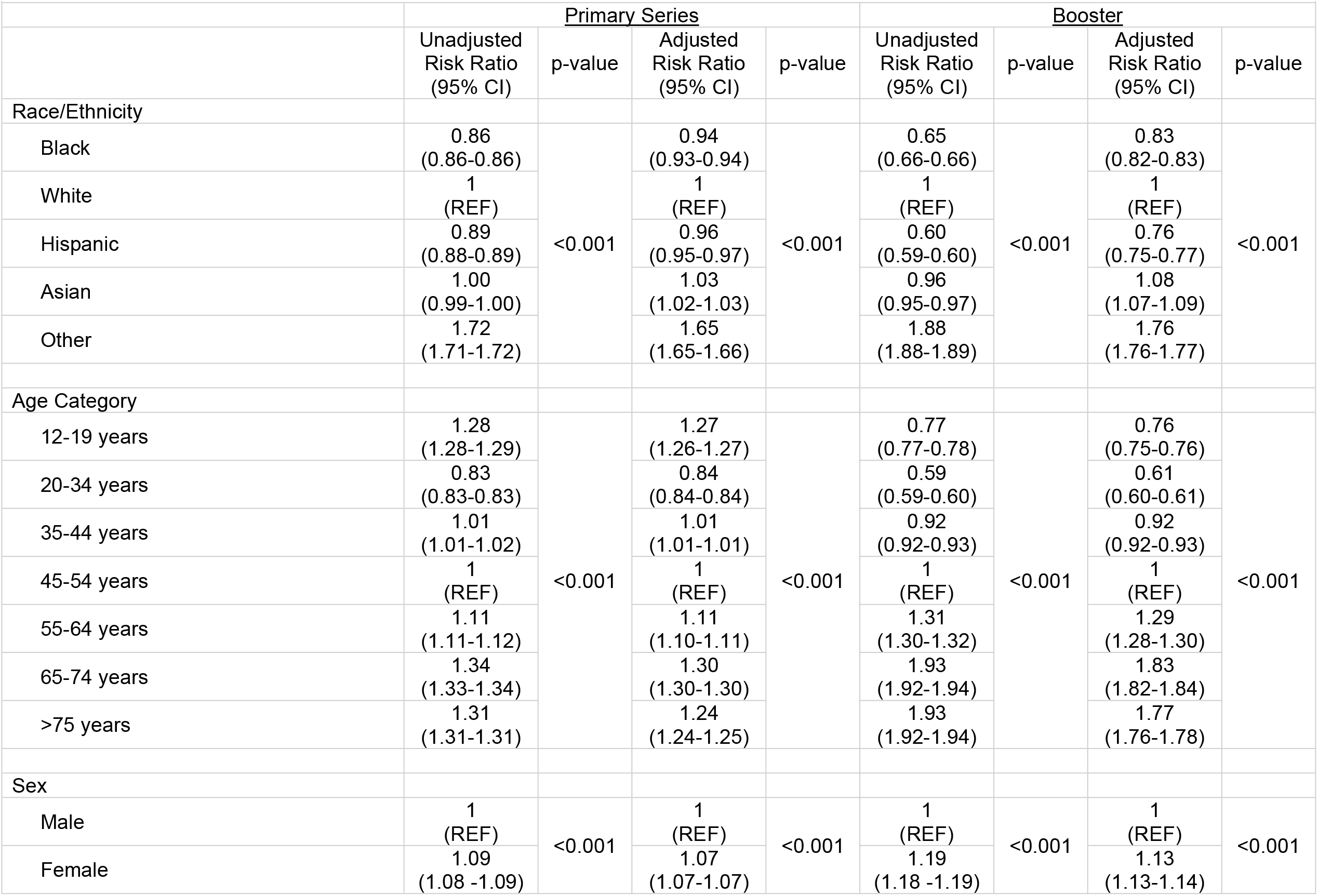
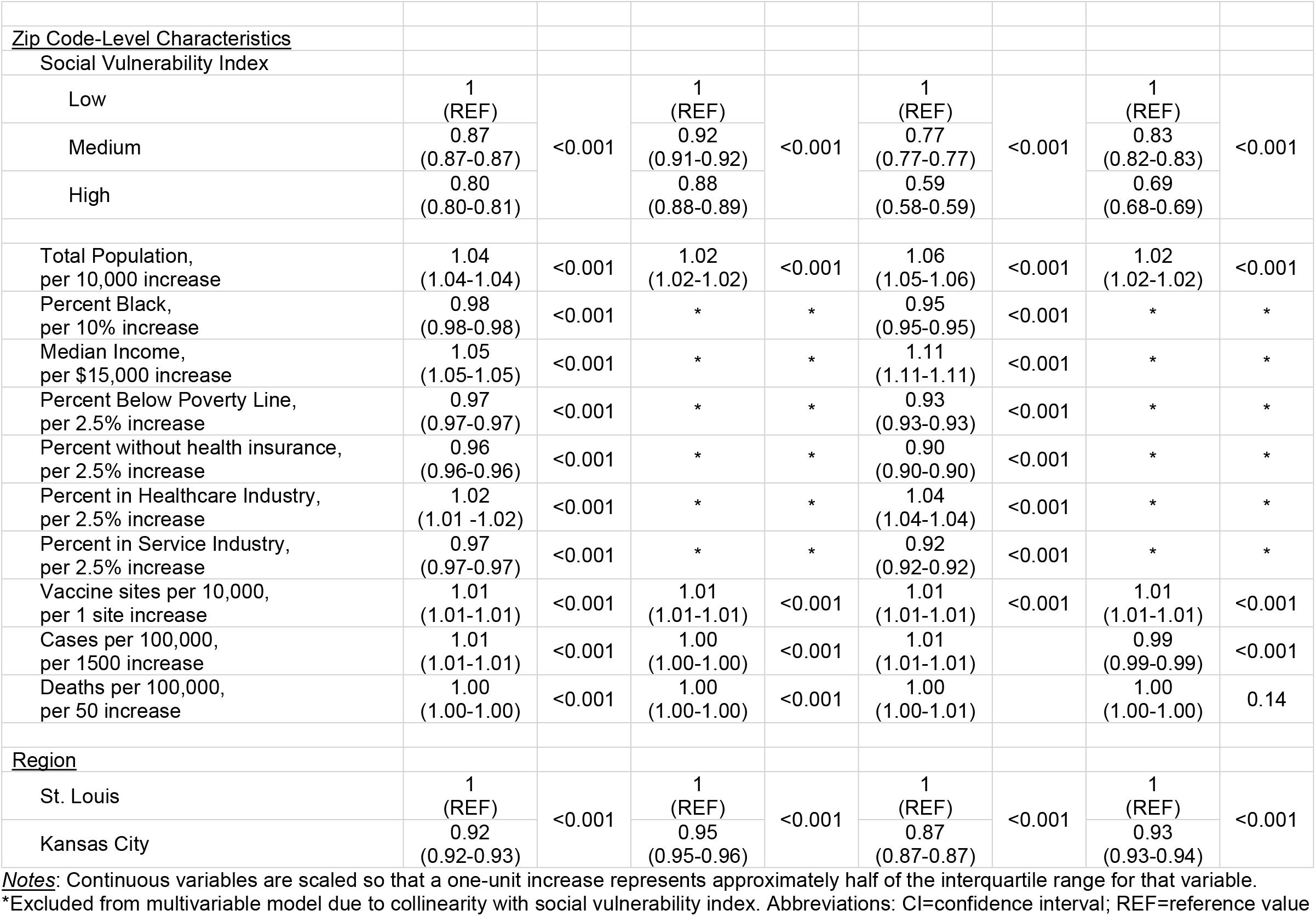
Poisson Model of Individual- and Zip Code-Level Factors Associated with Receipt of Primary COVID-19 Vaccination Series and Booster.

|  | Primary Series |  |  |  | Booster |  |  |  |
| --- | --- | --- | --- | --- | --- | --- | --- | --- |
|  | Unadjusted Risk Ratio (95% CI) | p-value | Adjusted Risk Ratio (95% CI) | p-value | Unadjusted Risk Ratio (95% CI) | p-value | Adjusted Risk Ratio (95% CI) | p-value |
| Race/Ethnicity |  |  |  |  |  |  |  |  |
| Black | 0.86<br>(0.86-0.86) | <0.001 | 0.94<br>(0.93-0.94) | <0.001 | 0.65<br>(0.66-0.66) | <0.001 | 0.83<br>(0.82-0.83) | <0.001 |
| White | 1<br>(REF) |  | 1<br>(REF) |  | 1<br>(REF) |  | 1<br>(REF) |  |
| Hispanic | 0.89<br>(0.88-0.89) |  | 0.96<br>(0.95-0.97) |  | 0.60<br>(0.59-0.60) |  | 0.76<br>(0.75-0.77) |  |
| Asian | 1.00<br>(0.99-1.00) |  | 1.03<br>(1.02-1.03) |  | 0.96<br>(0.95-0.97) |  | 1.08<br>(1.07-1.09) |  |
| Other | 1.72<br>(1.71-1.72) |  | 1.65<br>(1.65-1.66) |  | 1.88<br>(1.88-1.89) |  | 1.76<br>(1.76-1.77) |  |
| Age Category |  |  |  |  |  |  |  |  |
| 12-19 years | 1.28<br>(1.28-1.29) | <0.001 | 1.27<br>(1.26-1.27) | <0.001 | 0.77<br>(0.77-0.78) | <0.001 | 0.76<br>(0.75-0.76) | <0.001 |
| 20-34 years | 0.83<br>(0.83-0.83) |  | 0.84<br>(0.84-0.84) |  | 0.59<br>(0.59-0.60) |  | 0.61<br>(0.60-0.61) |  |
| 35-44 years | 1.01<br>(1.01-1.02) |  | 1.01<br>(1.01-1.01) |  | 0.92<br>(0.92-0.93) |  | 0.92<br>(0.92-0.93) |  |
| 45-54 years | 1<br>(REF) |  | 1<br>(REF) |  | 1<br>(REF) |  | 1<br>(REF) |  |
| 55-64 years | 1.11<br>(1.11-1.12) |  | 1.11<br>(1.10-1.11) |  | 1.31<br>(1.30-1.32) |  | 1.29<br>(1.28-1.30) |  |
| 65-74 years | 1.34<br>(1.33-1.34) |  | 1.30<br>(1.30-1.30) |  | 1.93<br>(1.92-1.94) |  | 1.83<br>(1.82-1.84) |  |
| >75 years | 1.31<br>(1.31-1.31) |  | 1.24<br>(1.24-1.25) |  | 1.93<br>(1.92-1.94) |  | 1.77<br>(1.76-1.78) |  |
| Sex |  |  |  |  |  |  |  |  |
| Male | 1<br>(REF) | <0.001 | 1<br>(REF) | <0.001 | 1<br>(REF) | <0.001 | 1<br>(REF) | <0.001 |
| Female | 1.09<br>(1.08 -1.09) |  | 1.07<br>(1.07-1.07) |  | 1.19<br>(1.18 -1.19) |  | 1.13<br>(1.13-1.14) |  |
| <u>Zip Code-Level Characteristics</u> |  |  |  |  |  |  |  |  |
| Social Vulnerability Index |  |  |  |  |  |  |  |  |
| Low | 1<br>(REF) | <0.001 | 1<br>(REF) | <0.001 | 1<br>(REF) | <0.001 | 1<br>(REF) | <0.001 |
| Medium | 0.87<br>(0.87-0.87) |  | 0.92<br>(0.91-0.92) |  | 0.77<br>(0.77-0.77) |  | 0.83<br>(0.82-0.83) |  |
| High | 0.80<br>(0.80-0.81) |  | 0.88<br>(0.88-0.89) |  | 0.59<br>(0.58-0.59) |  | 0.69<br>(0.68-0.69) |  |
| Total Population,<br>per 10,000 increase | 1.04<br>(1.04-1.04) | <0.001 | 1.02<br>(1.02-1.02) | <0.001 | 1.06<br>(1.05-1.06) | <0.001 | 1.02<br>(1.02-1.02) | <0.001 |
| Percent Black,<br>per 10% increase | 0.98<br>(0.98-0.98) | <0.001 | * | * | 0.95<br>(0.95-0.95) | <0.001 | * | * |
| Median Income,<br>per \$15,000 increase | 1.05<br>(1.05-1.05) | <0.001 | * | * | 1.11<br>(1.11-1.11) | <0.001 | * | * |
| Percent Below Poverty Line,<br>per 2.5% increase | 0.97<br>(0.97-0.97) | <0.001 | * | * | 0.93<br>(0.93-0.93) | <0.001 | * | * |
| Percent without health insurance,<br>per 2.5% increase | 0.96<br>(0.96-0.96) | <0.001 | * | * | 0.90<br>(0.90-0.90) | <0.001 | * | * |
| Percent in Healthcare Industry,<br>per 2.5% increase | 1.02<br>(1.01 -1.02) | <0.001 | * | * | 1.04<br>(1.04-1.04) | <0.001 | * | * |
| Percent in Service Industry,<br>per 2.5% increase | 0.97<br>(0.97-0.97) | <0.001 | * | * | 0.92<br>(0.92-0.92) | <0.001 | * | * |
| Vaccine sites per 10,000,<br>per 1 site increase | 1.01<br>(1.01-1.01) | <0.001 | 1.01<br>(1.01-1.01) | <0.001 | 1.01<br>(1.01-1.01) | <0.001 | 1.01<br>(1.01-1.01) | <0.001 |
| Cases per 100,000,<br>per 1500 increase | 1.01<br>(1.01-1.01) | <0.001 | 1.00<br>(1.00-1.00) | <0.001 | 1.01<br>(1.01-1.01) |  | 0.99<br>(0.99-0.99) | <0.001 |
| Deaths per 100,000,<br>per 50 increase | 1.00<br>(1.00-1.00) | <0.001 | 1.00<br>(1.00-1.00) | <0.001 | 1.00<br>(1.00-1.01) |  | 1.00<br>(1.00-1.00) | 0.14 |
| <u>Region</u> |  |  |  |  |  |  |  |  |
| St. Louis | 1<br>(REF) | <0.001 | 1<br>(REF) | <0.001 | 1<br>(REF) | <0.001 | 1<br>(REF) | <0.001 |
| Kansas City | 0.92<br>(0.92-0.93) |  | 0.95<br>(0.95-0.96) |  | 0.87<br>(0.87-0.87) |  | 0.93<br>(0.93-0.94) |  |

Notes: Continuous variables are scaled so that a one-unit increase represents approximately half of the interquartile range for that variable.
\*Excluded from multivariable model due to collinearity with social vulnerability index. Abbreviations: CI=confidence interval; REF=reference value

## DISCUSSION

Our analyses characterized disparities in the COVID-19 vaccination campaign in the St. Louis and Kansas City regions across racial/ethnic communities, across levels of social vulnerability, over time, and across types of vaccine administrations sites. We describe changes in the rates of receiving the primary COVID-19 vaccination series and boosters across race/ethnicity and social vulnerability and highlight how these changes corresponded with shifts in the types of locations where vaccines individuals were being vaccinated. We also use Lorenz curves and Gini coefficients to quantify disparities in vaccinations with respect to population, COVID-19 related disease burden, and social vulnerability. Overall, these results provide a deeper characterization the systemic inequities in distribution of one of the most critical (and initially scarce) resources for controlling the COVID-19 pandemic but one that is immediately actionable: COVID-19 vaccinations.

These analyses provide a deeper understanding of the patterns of vaccine inequities over time, and note that disparities were greatest earlier on but have also largely persisted over time with minimal improvement since April 2020. Furthermore, they emerged anew with the booster rollout in Fall 2021. Early during vaccination, rates of completing the primary vaccine series were highest among White and Asian individuals in zip codes with low SVIs. During this early period a vast majority of vaccines were administered through health systems and also mass vaccination sites coordinated by public health departments. The relationship between race/ethnicity and zip code SVI is salient during this period: Black and Hispanic individuals living in high SVI zip codes had strikingly lower rates of vaccination compared to other groups, whereas Black and Hispanic individuals in low SVI zip codes had similar to somewhat higher rates compared to White individuals in medium and high SVI zip codes. Over time, and particularly after all adults became eligible for vaccination, rates of vaccinations among Black and Hispanic individuals across all SVI zip codes started to exceed those among White individuals. During these periods, sites of vaccine administration also diversified and shifted more towards pharmacies and other small community-based sites (and were much less likely to occur at very large facilities). When quantifying these disparities using Lorenz curves, we note that disparities in vaccinations were highest relative to population-level social vulnerability and deaths, but still evident—albeit reduced—even when considering vaccinations relative to the overall population and diagnosed COVID-19 cases. Lastly, when examining disparities within zip codes, we see consistently higher rates of vaccination among White individuals compared to Black individuals, with the starkest difference in high SVI zip codes. Unfortunately, despite the slow progress from the early periods in improving equity in completion of the primary vaccine series, the same patterns of disparities were repeated again during the booster rollout, and were often of greater magnitude.

It is critical to understand these trends in the context of the underlying structural driving forces and decisions leading to these vaccination patterns, both of which are relevant nationally and not specific to Missouri. First, the high levels of disparities seen in the earlier stages of the primary vaccine series and booster rollout likely reflects the fact that health care workers and older individuals were eligible for vaccination first, factors that are also associated with higher socioeconomic status and lower SVI [9,10]. Second, the early phases for the primary vaccine series occurred primarily at sites associated with large health systems. However, these are also the sites from which Black and Hispanic individuals—and particularly those from high SVI zip codes—were comparatively less likely to ultimately receive vaccinations, highlighting a critical issue related to vaccine access among racially and ethnically marginalized and socially vulnerable communities [27-32]. Although large health systems may have been more readily able to overcome logistic issues and provide the robust cold chain needed for mRNA vaccines, they have limited mandates and expertise for implementing large-scale public health initiatives. Even prior to the pandemic, the significant disparities in who accesses care at these health systems and who is outside of them were well-known [30,33]. Physical access, challenges with scheduling (particularly online), disparities in insurance, lack of community partnerships, and mistrust of large institutions that have largely neglected underserved communities often serve as salient barriers to care-seeking in large health systems for individuals from high SVI communities [27,30,34,35]. Vaccination campaigns are a public health strategy that requires broad reach into communities that large health systems did not have and were not designed for; thus, the strategies relying on these systems did not reach the most vulnerable populations essentially by design even though the vaccines themselves were freely available. These patterns seen in both the primary vaccine series and booster rollout were also mirrored in prior research from our group examining disparities in COVID-19 testing, and their origins can be traced back to many of the same root causes [4]. Ultimately, the repeated reliance on systems with a history of providing lower access to certain segments of the population is representative of how structural inequities also became embedded in vaccine rollout from its onset and serves as a precautionary tale, albeit one that has been told too many times before.

Overall vaccinations rates and patterns over time in Black and Hispanic populations and high SVI zip codes further underscores the deeply embedded systemic nature of racialized disparities and the highly intersectional nature of systemic racism and social vulnerability [1,27-30,33-35]. Even though several vaccination strategies sought to prioritize Black and Hispanic individuals living in high SVI zip codes given their high burden of disease earlier on [11,12,36-38], they still had dramatically lower vaccination rates compared to White and Asian individuals in the same high SVI zip codes and those from zip codes with low SVIs. As the initial vaccine rollout progressed, though, rates in Black and Hispanic populations did eventually exceed those in White (though not Asian) populations. This coincided with wider vaccine availability and the shift toward vaccine administration at smaller centers such as pharmacies. Again, these changes in vaccination rates over time may be indicative of the increased access to vaccinations in Black, Hispanic, and other socially vulnerable communities through community-based settings as opposed to large health systems [30,34,37,39]. These patterns must also be contextualized within the growing literature on vaccine confidence and hesitancy.

Vaccine hesitancy is not monolithic and ranges from beliefs in conspiracy theories and skepticism about COVID-19 to more nuanced concerns regarding safety, side effects, inability to take time off work, observing others safely vaccinated (i.e., social proof), and lack of trusted messaging [29,33-35,40-42]; its patterns and trends across communities also varies [43,44]. Qualitative studies have shown that lack of vaccine confidence in Black communities in particular stems largely from histories of systematic mistreatment and racism—which includes failed contemporary responses to COVID-19—leading to mistrust of larger institutions and concerns over bearing the burden of unfavorable safety and side effect profiles (particularly given the rapid timeline of development and shifting messaging over the need for additional doses) [29,35]. However, rates of primary series completion in the Black population also likely increased as confidence in vaccinations improved over time, more of the population was safely vaccinated (i.e., social proof), purposeful and targeted messaging was delivered from trusted sources, and there were more opportunities to discuss specific questions and concerns with trusted health care providers [43,44]. Although a common pattern with the diffusion of many innovations, it is critical to contextualize the structural disparities leading to this late adoption.

Although multiple strategies were put forth early in order prioritize equitable vaccination, our analysis shows that we were far from achieving such goals when examined from several metrics. Early vaccine allocation strategies designed to maximize benefits when supply was limited included considerations for prioritizing groups with higher risk for COVID-19 exposure or who had experienced higher burden of COVID-19 disease using metrics such as geography, social vulnerability index, or race/ethnicity (in addition to using age, comorbidities, high risk occupations) [11-13,36-38]. Still, these strategies mostly focused on determining vaccine eligibility, but eligibility or availability of vaccines doesn’t equate to adequate access. Indeed, achieving equity would have also required early concomitant prioritization and efforts to target structural barriers to vaccine uptake and reasons for later adoption [45]. Several programs demonstrated success using early, low barrier, and widely available access to vaccines at community-based sites (as opposed to mass vaccination sites and large health systems, often requiring online registration) in areas with high social vulnerability coupled with abundant opportunities to connect with and discuss concerns with trusted sources of information [30,34,41,46-50]. A program in San Francisco leveraged a community-based vaccination site near a transportation hub to target both access and trust-related barriers, and leveraged both high-touch (e.g., going door-to-door to provide information and register individuals) and low-touch methods (e.g., flyers and advertisements [50]. Approaches like these are even more important during the later stages of vaccination roll-out, when large or mass vaccination sites—which allowed for high volume for those already eager to be vaccinated—are likely at the limits of their reach.

There are several limitations to our analysis. First, reporting of all vaccinations was mandated by the state, but race/ethnicity and zip code were not reported consistently, particularly at smaller sites. Still, as this missingness was highly dependent on the vaccination date and site date, multiple imputation would still yield unbiased results even with higher levels of missingness [24-26]. Second, there may also have been misclassification of zip codes of individuals if permanent addresses did not match where people were actually living at the time of vaccination or in our categorization of vaccine location types. However, any misclassification was likely small and there is no reason to believe that there was systemic error that would substantially bias our results. Third, we used zip code population estimates from the 2020 census data, but true population sizes—and thus the appropriate denominators for some analyses—may have changed since then, particularly due to the well-documented migrations that occurred during the early phases of the pandemic. Fourth, we lacked complementary data that could help contextualize our findings (e.g., association between race/ethnicity and time or vaccine location type) and help characterize the relationship with potential drivers of these disparities, such as occupation, health insurance status, linkage to primary care, and vaccination awareness, knowledge, beliefs, and intentions. Fourth, in this analysis we were unable to provide more granular details on other racial/ethnic minorities such as indigenous or multi-racial individuals, either due to small populations in the regions or inability to link these population across data sources. Still, although we do include Black, White, Hispanic, and Asian communities, it remains critical to also assess disparities across other minoritized communities, acknowledging that the multidimensional nature of health disparities and unique drivers across these different communities warrants dedicated attention and public health action.

In conclusion, we provide nuanced characterizations of the disparities in COVID-19 vaccination across racial/ethnic communities, across levels of social vulnerability, over time, and across types of vaccine administration sites after one year of vaccination. Equitable COVID-19 vaccination is one of the most critical targets for successfully ending the pandemic, but, despite substantial discussion on how to effectively do so, it is clear that our strategies—both nationally and in Missouri—have yet to overcome the deeply entrenched systemic inequities in health care and society. Future planning for proactive and considered public health strategies in the face of pandemic emergencies—as opposed to reactive approaches—are needed to ensure that our responses are equitable from the outset and do not disproportionately affect minority communities both in the United States and globally.

## Data Availability

The data underlying the results presented in the study are managed by the Missouri Department of Health and Senior Services and are not publicly available. Details on how to request data for public health research are available at https://health.mo.gov/data.

## REFERENCES

1. Bibbins-Domingo K. This Time Must Be Different: Disparities During the COVID-19 Pandemic. Ann Intern Med. 2020;173(3):233–4.

2. Mackey K, Ayers CK, Kondo KK, Saha S, Advani SM, Young S, et al. Racial and Ethnic Disparities in COVID-19–Related Infections, Hospitalizations, and Deaths. Annals of Internal Medicine. 2020;174(3):362–73.

3. Miller S, Wherry LR, Mazumder B. Estimated Mortality Increases During The COVID-19 Pandemic By Socioeconomic Status, Race, And Ethnicity. Health Affairs. 2021;40(8):1252–60.

4. Mody A, Pfeifauf K, Bradley C, Fox B, Hlatshwayo MG, Ross W, et al. Understanding Drivers of Coronavirus Disease 2019 (COVID-19) Racial Disparities: A Population-Level Analysis of COVID-19 Testing Among Black and White Populations. Clin Infect Dis. 2021;73(9):e2921–e31.

5. Bajos N, Jusot F, Pailhé A, Spire A, Martin C, Meyer L, et al. When lockdown policies amplify social inequalities in COVID-19 infections: evidence from a cross-sectional population-based survey in France. BMC Public Health. 2021;21(1):705.

6. Zhou M, Kan M-Y. The varying impacts of COVID-19 and its related measures in the UK: A year in review. PLoS ONE. 2021;16(9):e0257286.

7. Galea S, Abdalla SM. COVID-19 Pandemic, Unemployment, and Civil Unrest: Underlying Deep Racial and Socioeconomic Divides. JAMA. 2020;324(3):227–8.

8. Benfer EA, Vlahov D, Long MY, Walker-Wells E, Pottenger JL, Jr., Gonsalves G, et al. Eviction, Health Inequity, and the Spread of COVID-19: Housing Policy as a Primary Pandemic Mitigation Strategy. J Urban Health. 2021;98(1):1–12.

9. National Academies of Sciences E, and Medicine. Framework for Equitable Allocation of COVID-19 Vaccine. Washington, DC: The National Academies Press; 2020.

10. Bell BP, Romero JR, Lee GM. Scientific and Ethical Principles Underlying Recommendations From the Advisory Committee on Immunization Practices for COVID-19 Vaccination Implementation. JAMA. 2020;324(20):2025–6.

11. Schmidt H, Weintraub R, Williams MA, Miller K, Buttenheim A, Sadecki E, et al. Equitable allocation of COVID-19 vaccines in the United States. Nature Medicine. 2021;27(7):1298–307.

12. Wrigley-Field E, Kiang MV, Riley AR, Barbieri M, Chen Y-H, Duchowny KA, et al. Geographically targeted COVID-19 vaccination is more equitable and averts more deaths than age-based thresholds alone. Science Advances. 2021;7(40):eabj2099.

13. Srivastava T, Schmidt H, Sadecki E, Kornides ML. Disadvantage Indices Deployed to Promote Equitable Allocation of COVID-19 Vaccines in the US: A Scoping Review of Differences and Similarities in Design. JAMA Health Forum. 2022;3(1):e214501–e.

14. Lorenz MO. Methods of Measuring the Concentration of Wealth. Publications of the American Statistical Association. 1905;9(70):209–19.

15. Centers for Disease Control and Prevention/ Agency for Toxic Substances and Disease Registry/ Geospatial Research A, and Services Program. CDC/ATSDR Social Vulnerability Index 2018 Database [Available from: https://www.atsdr.cdc.gov/placeandhealth/svi/data_documentation_download.html.

16. Christopoulos KA, Hartogensis W, Glidden DV, Pilcher CD, Gandhi M, Geng EH. The Lorenz curve: a novel method for understanding viral load distribution at the population level. AIDS. 2017;31(2):309–10.

17. Mauguen A, Begg CB. Using the Lorenz Curve to Characterize Risk Predictiveness and Etiologic Heterogeneity. Epidemiology. 2016;27(4):531–7.

18. Rita Neves Costa SbPr-D. Statistics Paper Series: Not all inequality measures were created equal. ECB Statistics Paper Series. December 2019;31.

19. Zou G. A modified poisson regression approach to prospective studies with binary data. Am J Epidemiol. 2004;159(7):702–6.

20. Zou GY, Donner A. Extension of the modified Poisson regression model to prospective studies with correlated binary data. Stat Methods Med Res. 2013;22(6):661–70.

21. Jones CP. Invited commentary: “race,” racism, and the practice of epidemiology. Am J Epidemiol. 2001;154(4):299–304; discussion 5-6.

22. Chowkwanyun M, Reed AL, Jr. Racial Health Disparities and Covid-19 - Caution and Context. N Engl J Med. 2020;383(3):201–3.

23. Boyd RW, Lindo EG, Weeks LD, McLemore MR. On Racism: A New Standard For Publishing On Racial Health Inequities [Internet]: Health Affairs Blog. 2020.

24. Lee KJ, Carlin JB. Multiple imputation for missing data: fully conditional specification versus multivariate normal imputation. Am J Epidemiol. 2010;171(5):624–32.

25. Madley-Dowd P, Hughes R, Tilling K, Heron J. The proportion of missing data should not be used to guide decisions on multiple imputation. J Clin Epidemiol. 2019;110:63–73.

26. Bernaards CA, Belin TR, Schafer JL. Robustness of a multivariate normal approximation for imputation of incomplete binary data. Stat Med. 2007;26(6):1368–82.

27. Jean-Jacques M, Bauchner H. Vaccine Distribution—Equity Left Behind? JAMA. 2021;325(9):829–30.

28. Thakore N, Khazanchi R, Orav EJ, Ganguli I. Association of Social Vulnerability, COVID-19 vaccine site density, and vaccination rates in the United States. Healthcare. 2021;9(4):100583.

29. Balasuriya L, Santilli A, Morone J, Ainooson J, Roy B, Njoku A, et al. COVID-19 Vaccine Acceptance and Access Among Black and Latinx Communities. JAMA Network Open. 2021;4(10):e2128575–e.

30. Njoku A, Joseph M, Felix R. Changing the Narrative: Structural Barriers and Racial and Ethnic Inequities in COVID-19 Vaccination. International journal of environmental research and public health. 2021;18(18):9904.

31. Blackstock U, Blackstock O. White Americans are being vaccinated at higher rates than Black Americans. Such inequity cannot stand.: Washington Post; February 1, 2021 [Available from: https://www.washingtonpost.com/opinions/2021/02/01/racial-inequality-covid-vaccine/.

32. Boyd R. Black People Need Better Vaccine Access, Not Better Vaccine Attitudes: New York Times; March 5, 2021 [Available from: https://www.nytimes.com/2021/03/05/opinion/us-covid-black-people.html.

33. Levesque JF, Harris MF, Russell G. Patient-centred access to health care: conceptualising access at the interface of health systems and populations. Int J Equity Health. 2013;12:18.

34. Marcelin JR, Swartz TH, Bernice F, Berthaud V, Christian R, da Costa C, et al. Addressing and Inspiring Vaccine Confidence in Black, Indigenous, and People of Color During the Coronavirus Disease 2019 Pandemic. Open forum infectious diseases. 2021;8(9):ofab417–ofab.

35. Momplaisir F, Haynes N, Nkwihoreze H, Nelson M, Werner RM, Jemmott J. Understanding Drivers of COVID-19 Vaccine Hesitancy Among Blacks. Clin Infect Dis. 2021.

36. Ndugga N, Artiga S, Pham O. How are States Addressing Racial Equity in COVID-19 Vaccine Efforts? : Kaiser Family Foundation; March 10, 2021 [Available from: https://www.kff.org/racial-equity-and-health-policy/issue-brief/how-are-states-addressing-racial-equity-in-covid-19-vaccine-efforts/.

37. Bibbins-Domingo K, Petersen M, Havlir D. Taking Vaccine to Where the Virus Is—Equity and Effectiveness in Coronavirus Vaccinations. JAMA Health Forum. 2021;2(2):e210213–e.

38. Schmidt H, Gostin LO, Williams MA. Is It Lawful and Ethical to Prioritize Racial Minorities for COVID-19 Vaccines? JAMA. 2020;324(20):2023–4.

39. Corallo B, Artiga S, Tolbert J. Are Health Centers Facilitating Equitable Access to COVID-19 Vaccinations? : Kaiser Family Foundation; June 02, 2021 [Available from: https://www.kff.org/coronavirus-covid-19/issue-brief/are-health-centers-facilitating-equitable-access-to-covid-19-vaccinations-a-june-2021-update/.

40. Tram KH, Saeed S, Bradley C, Fox B, Eshun-Wilson I, Mody A, et al. Deliberation, Dissent, and Distrust: Understanding distinct drivers of COVID-19 vaccine hesitancy in the United States. Clin Infect Dis. 2021.

41. Corbie-Smith G. Vaccine Hesitancy Is a Scapegoat for Structural Racism. JAMA Health Forum. 2021;2(3):e210434–e.

42. Eshun-Wilson I, Mody A, Tram KH, Bradley C, Sheve A, Fox B, et al. Preferences for COVID-19 vaccine distribution strategies in the US: A discrete choice survey. PLoS ONE. 2021;16(8):e0256394–e.

43. Padamsee TJ, Bond RM, Dixon GN, Hovick SR, Na K, Nisbet EC, et al. Changes in COVID-19 Vaccine Hesitancy Among Black and White Individuals in the US. JAMA Netw Open. 2022;5(1):e2144470.

44. Daly M, Jones A, Robinson E. Public Trust and Willingness to Vaccinate Against COVID-19 in the US From October 14, 2020, to March 29, 2021. JAMA. 2021;325(23):2397–9.

45. Hardeman A, Wong T, Denson JL, Postelnicu R, Rojas JC. Evaluation of Health Equity in COVID-19 Vaccine Distribution Plans in the United States. JAMA Network Open. 2021;4(7):e2115653–e.

46. Abdul-Mutakabbir JC, Casey S, Jews V, King A, Simmons K, Hogue MD, et al. A three-tiered approach to address barriers to COVID-19 vaccine delivery in the Black community. The Lancet Global health. 2021;9(6):e749–e50.

47. Galaviz KI, Breland JY, Sanders M, Breathett K, Cerezo A, Gil O, et al. Implementation Science to Address Health Disparities During the Coronavirus Pandemic. Health Equity. 2020;4(1):463–7.

48. Faherty LJ, Ringel JS, Williams MV, Kranz AM, Perez L, Schulson L, et al. Early Insights from the Equity-First Vaccination Initiative. Santa Monica, CA: RAND Corporation; 2021.

49. Beste Lauren A, Chen A, Geyer J, Wilson M, Schuttner L, Wheat C, et al. Best Practices for an Equitable Covid-19 Vaccination Program. NEJM Catalyst. 2(10).

50. Marquez C, Kerkhoff AD, Naso J, Contreras MG, Castellanos Diaz E, Rojas S, et al. A multi-component, community-based strategy to facilitate COVID-19 vaccine uptake among Latinx populations: From theory to practice. PLoS ONE. 2021;16(9):e0257111–e.

